# Immunity to SARS-CoV-2 persists 9 months post-symptoms with an altered T cell phenotype compared to influenza A virus-specific memory

**DOI:** 10.1101/2021.06.08.21258518

**Authors:** Jaclyn C. Law, Melanie Girard, Gary Y. C. Chao, Lesley A. Ward, Baweleta Isho, Bhavisha Rathod, Karen Colwill, Zhijie Li, James M. Rini, Feng Yun Yue, Samira Mubareka, Allison J. McGeer, Mario A. Ostrowski, Jennifer L. Gommerman, Anne-Claude Gingras, Tania H. Watts

## Abstract

SARS-CoV-2 induces T cell, B cell and antibody responses that are detected for several months in recovered individuals. Whether this response resembles a typical respiratory viral infection is a matter of debate. Here we followed T cell and antibody responses in 24 mainly non-hospitalized SARS-CoV-2 recovered subjects at two time points (median of 45- and 145-days post-symptom onset). Antibody responses were detected in 95% of subjects, with a strong correlation between plasma and salivary anti-S and anti-RBD IgG, as well as a correlation between circulating T follicular helper cells and the SARS-CoV-2-specific IgG response. Based on intracellular cytokine production or proliferation, CD4^+^ T cell responses to SARS-CoV-2 were detected in all subjects, decaying with a half-life of 5-6 months for S-specific IL-2-producing cells. CD4^+^ responses were largely of the T helper 1 phenotype, but with a lower ratio of IFN-γ: IL-2 producing cells and a lower frequency of CD8^+:^ CD4^+^ T cells compared to influenza A virus-(IAV)-specific memory responses within the same subjects. Analysis of secreted molecules also revealed a lower ratio of IFN-γ: IL-2 and IFN-γ: IL-6 and an altered cytotoxic profile for S- and N-specific compared to IAV-specific responses. These data suggest that the memory T-cell phenotype after a single infection with SARS-CoV-2 persists over time, with an altered cytokine and cytotoxic profile compared to long term memory to IAV within the same subjects.

**One Sentence Summary:** Immunity to SARS-CoV-2 in a cohort of patients, mainly with mild COVID-19 disease, persists to 9 months with an altered T cell cytokine and cytotoxicity profile compared to influenza A virus-specific memory T cells from the same subjects.

## Introduction

SARS-CoV-2, a pandemic respiratory virus, continues to circulate in many regions around the world (https://coronavirus.jhu.edu/map.html). Immunity to SARS-CoV-2 in a high proportion of the population will be required to control this pandemic (*1*). Although vaccine-induced immunity is an important component of protection against SARS-CoV-2, a substantial number of people have recovered from COVID-19 and there is a need to understand the persistence and quality of immunity to SARS-CoV-2 in these individuals. All successful viruses must suppress the host innate immune response to some extent, however, SARS-CoV-2 is particularly adept at evading type I and III interferon (IFN) responses (*2–5*) and people with defects in IFN signaling are over-represented among severe cases (*6, 7*). Whether these unique features of the early response to SARS-CoV-2 impact long-term immunity to SARS-CoV-2 compared to other respiratory viruses remains unknown.

There is now extensive evidence that CD4^+^ and CD8^+^ T cell as well as antibody responses are induced following asymptomatic, mild or severe COVID19 infection, with memory T cell responses established within a few weeks of infection (*8–25*). CD4^+^ T cell responses to SARS-CoV-2 are largely of the Th1 type, with cytotoxic CD4^+^ T cells and circulating T follicular helper cells (cTfh) also detected in some studies (*8, 11, 16, 19*). CD8^+^ T cells are less consistently detected (*8, 11, 16, 19*). Spike (S), Nucleocapsid (N), Membrane (M) and *Open reading frame* (*Orf*)3 are all major sources of CD4^+^ and CD8^+^ T cell epitopes (*8, 11, 16, 18, 20, 21, 26*). SARS-CoV-2 infection can lead to mild or severe outcomes with early T cell responses, increased frequency of airway T cells as well as early S-specific neutralizing and non-neutralizing antibody responses correlating with earlier viral control and better outcomes (*16, 17, 20, 27-33*). Several studies have demonstrated the persistence of T cell and/or antibody responses to SARS-CoV-2 out to 6-10 months post-infection (*34–40*). For example, Dan et al. reported that T cell responses to SARS-CoV-2 decline with a half-life of 3-5 months, with B cell memory and antibody responses relatively stable over 6 months (*34*).

In a previous study, we examined the memory response to SARS-CoV-2 in the early convalescent phase in a cohort of 13 subjects, 46% of whom were categorized as having moderate to severe disease based on admission to hospital or an intensive care unit (ICU), respectively (*19*). We compared T cell responses to SARS CoV-2 antigens (Ags) with memory responses to H1N1 IAV within the same subjects. At the early convalescent stage, CD4^+^ T cell responses predominated over CD8^+^ T cell responses, in contrast to the response to IAV, which was dominated by a stronger CD8^+^ T cell response (*19*). In addition, we noted a lower frequency of IFN-γ compared to IL-2 producing S-specific CD4^+^ T cells compared to IAV-specific responses which were dominated by IFN-γ specific responses (*19*), albeit these findings were limited by small sample size and potentially impacted by the predominance of severe cases in the cohort. Here we examined the persistence and phenotype of T cell responses to SARS-CoV-2 at two time points over a period of 9 months post-symptom onset (PSO) in a cohort of 24 confirmed COVID-19 convalescent subjects, 75% of whom were not hospitalized. The results show that even after mild COVID-19, T cell recall responses exhibit a lower CD8^+^: CD4^+^ T cell ratio and a higher proportion of IL-2 and IL-6 producing cells, as well as an altered cytotoxic profile compared to IAV-specific memory responses within the same subjects. These T cell responses and altered phenotype persist to about 9 months post-symptoms, with a half-life of 5-6 months for S-specific CD4^+^IL-2^+^ T cells. Plasma IgG responses to SARS-CoV-2 declined only slightly over the course of the study, whereas salivary antibody responses fell off more rapidly, albeit plasma and salivary IgG responses were strongly correlated overall.

## Results

### Study subjects

24 COVID-19 convalescent subjects, who had recovered from COVID-19 as confirmed by positive nasopharyngeal COVID-19 PCR upon presentation, consented to blood draw and saliva sampling. Samples were collected at two time points, the first between 30-154 days PSO, median 45 days, and the second between 55-249 days PSO, median 145 days. 18 subjects had mild disease (not hospitalized), 1 had moderate disease (hospitalized, non-ICU) and 5 subjects had severe disease (ICU). The median age was 46.5 years, and 50% were female (**Table S1**).

### Intracellular cytokine production by SARS-CoV-2 or IAV-specific CD4^+^ T cells over time

To assess the recall response to SARS-CoV-2 in convalescent subjects, we used 15-mer peptide pools with 11 amino acids overlap, comprising the S, M, N and Envelope (E) of the SARS-CoV-2 virus, to stimulate freshly thawed PBMC. Due to the large size of S, two S peptide pools, S1 and S2 pools were analyzed separately to reduce competition for Ag presentation. After 18h of stimulation, SARS-CoV-2 specific CD4^+^ T cells were detected based on the production of IL-2, IFN-γ, or TNF by intracellular cytokine staining (ICC) as determined by the gating strategy shown in **Fig S1A**. As most adults are expected to have a memory response to IAV, we used IAV strain PR8/34/H1N1 as an internal positive control to ensure the ability of each patient sample to mount a T cell recall response; phorbol myristic acetate/ionomycin (P/I) was used as a secondary non-specific stimulus to validate overall PBMC quality. Only samples with a response to P/I were included in the study and each response was considered positive if cytokine production was 10% over background. IL-2 producing CD4^+^ T cells were the most abundantly detected of the cytokine-producing CD4^+^ T cells in response to SARS-CoV-2 peptide pools, with 83% of donors at their first visit responding to S1, 88% to N and 88% to IAV (**Fig. 1A**). IL-2 responses persisted for at least 35 weeks, decaying at approximately 1.7%/week in response to S1 (t1/2=29 weeks) whereas the decay of the T cell response to IAV was more gradual at about 1%/week (t1/2 =50 weeks) (**Fig. 1A**).

**Fig. 1.**
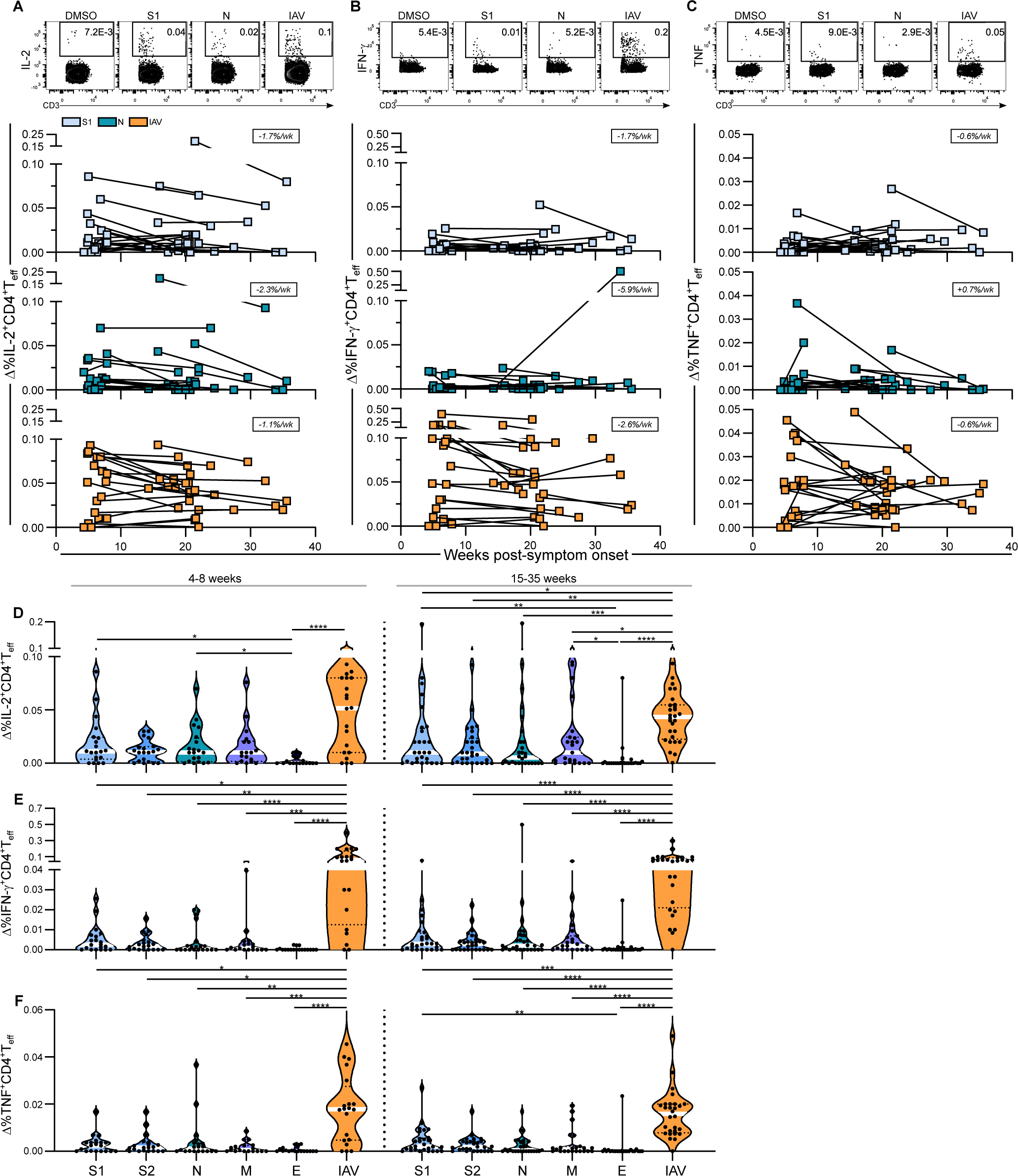
Intracellular cytokine production by SARS-CoV-2 or IAV-specific CD4^+^ T cells over time. Cytokine production by virus-specific CD4^+^ T cells was assessed by flow cytometry following 18h of stimulation with SARS-CoV-2 peptide pools or IAV. Representative flow cytometry plots (top) and longitudinal analyses of background subtracted values (Δ) show the frequency of CD4^+^ T cells expressing **(A)** IL-2, **(B)** IFN-γ and **(C)** TNF. Boxed numbers in each panel show the average rate of decay between time points for each response, calculated as described in the data analysis section. Violin plots show the median frequency of CD4^+^ T cells expressing **(D)** IL-2, **(E)** IFN-γ and **(F)** TNF to each antigen binned by time PSO (4-8 weeks or 15-35 weeks). All plots show data from 24 donors. Comparisons between each antigen were made using Dunn’s multiple comparisons test. Wk = week. *P≤0.05, **P≤0.01, ***P≤0.001, ****P≤0.0001.

IFN-γ producing CD4^+^ cells were detected at a lower frequency than IL-2 producing CD4^+^ T cells in response to SARS-CoV-2 peptide pools, with 79% of donors showing IFN-γ production in response to S1 and 67% in response to N, whereas 92% of donors had a response to IAV (**Fig. 1B**). TNF-producing cells were the lowest in frequency compared to IL-2 or IFN-γ producing cells (**Fig. 1C**). IL-2, IFN-γ and TNF-producing cells were also detected in response to S2, M and E peptide pools, and showed rates of decay similar to responses against S1 (**Fig. S1B-D**). Of the 5 SARS-CoV-2 peptide pools tested, the highest frequency of CD4^+^ T cell responses was detected against S, and the lowest frequency of responses was detected against E based on the 3 cytokines measured (**Fig. 1D-F**). In general, donors who had the strongest responses to S1 also had the strongest responses to all of the peptide pools tested. The response to P/I over time declined at a similar rate to the response to IAV, suggesting that overall T cell memory was declining slowly over this period (**Fig. S1B-D**). This was not due to a change in the frequency of Ag presenting cells between the two timepoints, as there was no difference in the frequency of HLA-DR^+^CD3^-^ cells over time (**Fig. S1E**). Altogether, the data show that CD4^+^ T cell responses against at least one SARS-CoV-2 Ag are detected in most donors. On average, SARS-CoV-2 specific T cells declined with an estimated half-life of 25 weeks (**Fig. 1 and S1**), whereas the memory response to IAV is more stable, with a half-life of closer to 1 year. Responses against IAV were generally higher than those against SARS-CoV-2 peptide pools, likely due to the use of whole IAV, containing the full complement of epitopes, as compared to individual peptide pools for SARS-CoV-2, as well as a lifetime of exposure to IAV or influenza vaccines in these adult subjects compared to a first response to SARS-CoV-2.

### Immunophenotype of SARS-CoV-2 and IAV-specific CD4^+^ T cell responses over time

We next characterized the phenotype of the SARS-CoV-2 specific CD4^+^ T cells in comparison to IAV-specific CD4^+^ T cells for production of 1,2 or 3 cytokines per cell as well as surface marker upregulation. Eighty-four percent of S-specific CD4^+^ T cells produced only 1 cytokine at 4-8 weeks PSO and this was similar at 15-35 weeks (**Fig 2A**). IAV-specific CD4^+^ T cell responses were slightly more multifunctional, with 4.6% of IAV-specific CD4^+^ T cells producing 3 cytokines at 4-8 weeks PSO and 5.7% at 15-35 weeks PSO. This was significantly greater than S1-specific CD4^+^ T cell responses, where only 2% of cells produced 3 cytokines at 4-8 weeks PSO, and 3.2% at 15-35 weeks PSO (**Fig. 2A**). The ratio of IFN-γ: IL-2 producing CD4^+^ T cells was also significantly lower in response to both S1 and N compared to IAV at 4-8 weeks PSO within the same donor (**Fig. 2B**). Moreover, this diminished IFN-γ:IL-2 ratio in the response to the S1 peptide pool compared to IAV persisted to 35 weeks (**Fig. 2B**).

**Fig. 2.**
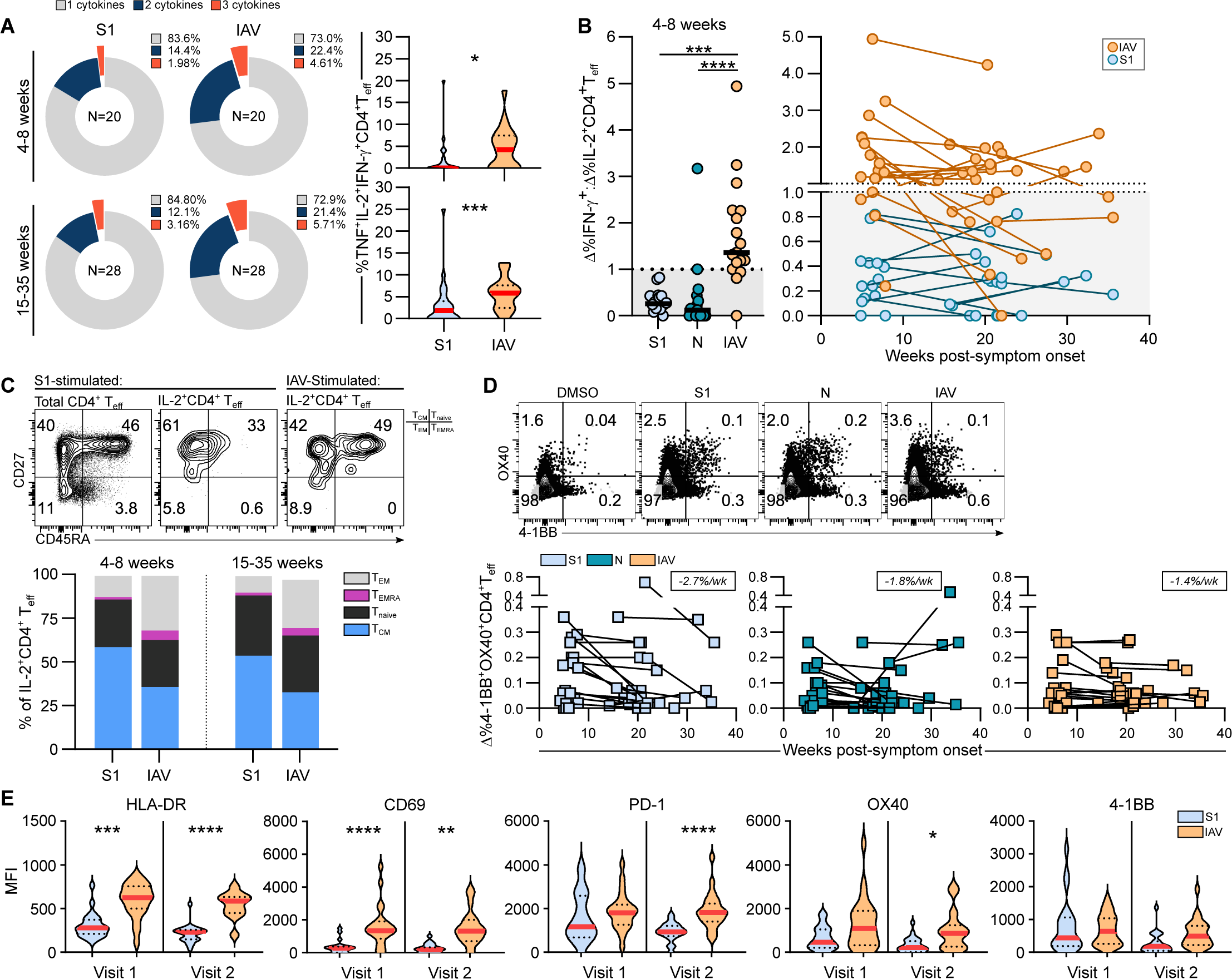
Immunophenotype of SARS-CoV-2 and IAV-specific CD4^+^ T cell responses over time. **(A)** Frequency of CD4^+^ T cells expressing IL-2, IFN-γ, and/or TNF as a proportion of cytokine-producing cells. **(B)** Ratio of %IFN-γ^+^: %TNF^+^ CD4^+^ T cells in donors producing both cytokines in response to S, N or IAV, showing comparison between S, N and IAV at 4-8 weeks (left) and the ratio over time up to 35 weeks (right), N=24. **(C)** Representative flow plot of CD45RA and CD27 expression by IL-2 producing CD4^+^ T cells in response to stimulation with S1 or IAV (top). Bar graph depicts each memory subset as a proportion of IL-2^+^CD4^+^ T cells (bottom), N=20. **(D)** Representative flow plots of 4-1BB and OX40 expression by CD4^+^ T cells in response to stimulation with S1, N or IAV. Graphs depict the %4-1BB^+^OX40^+^ CD4^+^ T cells over time, N=24. Boxed numbers in each panel show the average rate of decay between time points. **(E)** Expression of the indicated activation markers by IL-2^+^CD4^+^ T cells based on MFI, N=20. Violin plots show median (red line) and quartiles (dotted black line). Comparisons were made by Dunn’s multiple comparisons test. Wk = week. *P≤0.05, **P≤0.01, ***P≤0.001, ****P≤0.0001.

Based on the expression of CD27 and CD45RA by the IL-2 producing CD4^+^ T cells, all 4 subtypes of memory T cells were detected, with the largest fraction of IAV-specific and S-specific CD4^+^ T cells comprising the central memory (Tcm) phenotype (CD45RA^-^CD27^+^) (**Fig. 2C**). Compared to IAV-specific CD4^+^ T cells, S-specific CD4^+^ T cells had a higher proportion of Tcm and lower proportions of effector memory (CD45RA^-^CD27^-^) and T effector memory RA^+^ (CD45RA^+^CD27) cells (Temra), thought to represent more differentiated effector memory cells (**Fig. 2C, S2**). Activation markers such as CD134 (OX40), CD137 (4-1BB), HLA-DR, or CD69, are often used to identify Ag-specific T cells. Based on co-expression of OX40 and 4-1BB, 92% of donor PBMC showed a response to S1 peptide pools, 92% responded to N, and 83% responded to IAV at the 1^st^ visit. The rate of decay of these responses was similar to that observed based on cytokine production with a half-life of 5-6 months for SARS-CoV-2 specific responses (**Fig. 2D**). The expression of HLA-DR and CD69 on S-specific T cells was significantly lower than on IAV-specific T cells. PD-1 and OX40 expression were also lower on S-specific T cells than IAV-specific cells at the 2^nd^ visit, whereas there was no difference in 4-1BB expression at either timepoint (**Fig. 2E**). Thus, although the IAV-specific and SARS-CoV-2-specific cells had a similar preponderance of central memory phenotype cells, SARS-CoV-2 S- and N-specific T cell frequency showed a lower IFN-γ to IL-2 ratio and the T cells were less multifunctional than IAV-specific T cells within the same subjects.

### CD8^+^ T cell responses to SARS-CoV-2 are low in frequency

Next, we analyzed CD8^+^ T cell responses in the PBMC cultures following stimulation with SARS-CoV-2 S, N, M and E peptide pools compared to IAV at each time point. In general, the frequency of SARS-CoV-2-specific CD8^+^ T cell responses detected based on IFN-γ production was quite low. SARS-CoV-2-specific CD8^+^ responses were detected in 54% of donors in response to S1, 33% to S2, 29% to N, 9% to M, and 17% to E. This contrasted with 92% detected in response to IAV. The decay of these responses over time was 1-2%/week (**Fig. 3A**). As with the CD4^+^ T cell responses, SARS-CoV-2-specific IFN-γ-producing CD8^+^ T cells were most readily detected in response to S1 peptide pools and were detected in less than half of the study subjects in response to N, M or E peptide pools (**Fig. 3B**). Based on the expression of CD27 and CD45RA by IFN-γ^+^CD8^+^ T cells, both IAV and SARS-CoV-2 recall responses were predominantly of the more terminally differentiated Temra phenotype and there was no difference in the distribution of memory subsets between S-specific and IAV-specific CD8^+^ T cells. (**Fig. 3C, S3**). With the limitation that we had only a few severe cases, disease severity did not appear to impact the strength of the memory response (**Fig. 3D**). In sum, CD8^+^ T cell responses are present at low frequencies in response to SARS-CoV-2 peptide pools based on ICC staining with the response to S predominating over the other Ags tested at both time points.

**Fig. 3.**
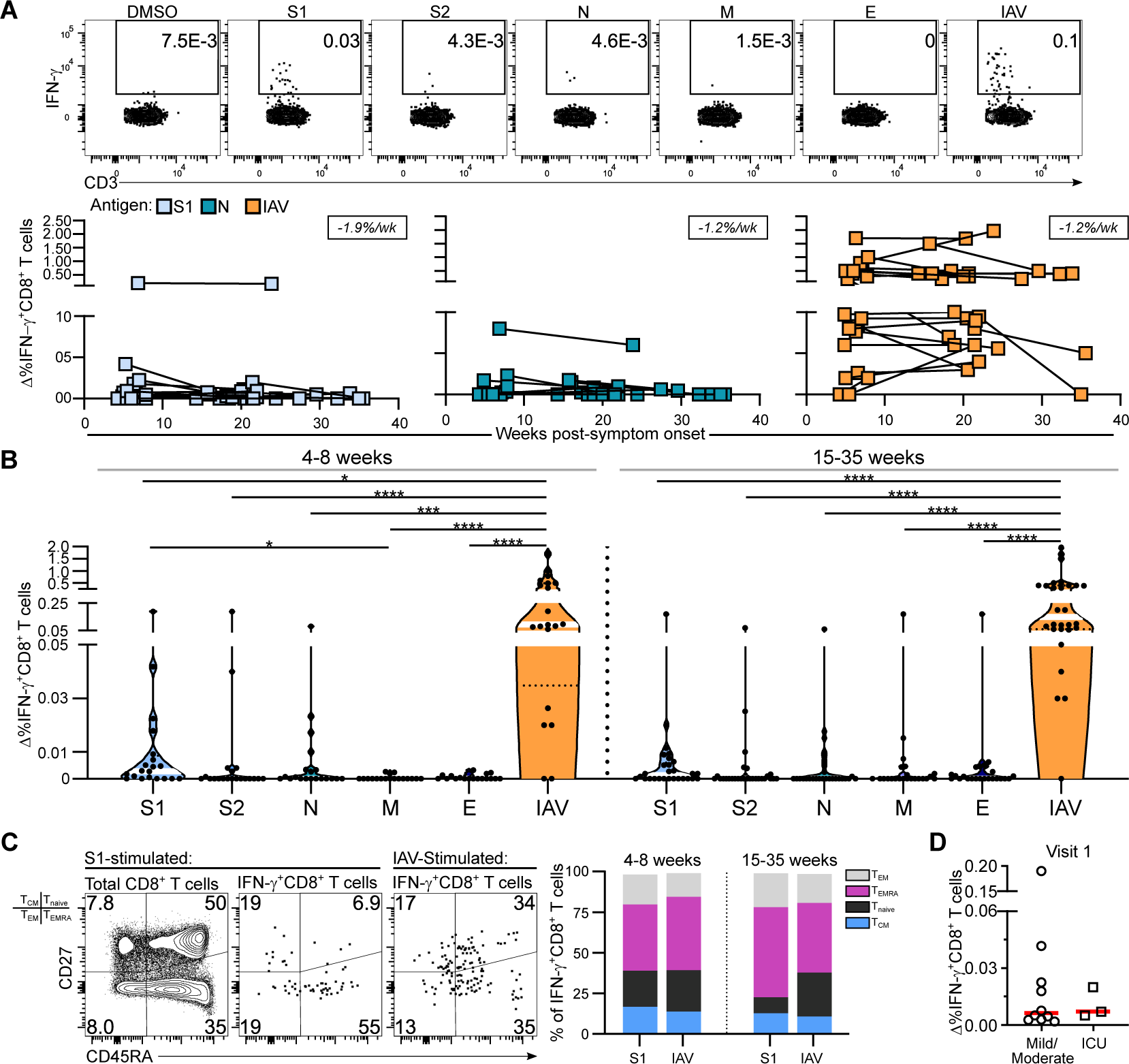
CD8^+^ T cell responses to SARS-CoV-2 are low in frequency. **(A)** Representative flow cytometry plots and longitudinal analyses of background subtracted values (Δ) show the frequency of CD8^+^ T cells expressing IFN-γ to the indicated antigen. Boxed numbers in each panel show the average rate of decay between time points for each response. **(B)** Violin plots show the median frequency (white line) of CD8^+^ T cells expressing IFN-γ to each antigen binned by time PSO (4-8 weeks or 15-35 weeks). **(A)** and **(B)** show data from 24 donors. Comparisons were made by Dunn’s multiple comparisons test. **(C)** Representative flow plot of CD45RA and CD27 expression by total and IFN-γ-producing CD8^+^ T cells (left). Bar graph depicts each memory subset as a proportion of IFN-γ^+^CD8^+^ T cells (right), N=13. **(D)** Comparison of the %IFN-γ^+^CD8^+^ T cells in mild/moderate vs ICU subjects by Mann-Whitney test. Wk = week. *P≤0.05, ***P≤0.001, ****P≤0.0001.

### Robust CD4^+^ and CD8^+^ T cell proliferative responses to SARS-CoV-2 up to 35 weeks PSO

As CD8^+^ T cell responses to SARS-CoV-2 were low in frequency as detected by overnight stimulation, we next asked if these cells could be better detected based on Ag-specific proliferation, an important indicator of potential recall responses. To this end, total PBMC were labelled with carboxyfluorescein succinimidyl ester (CFSE) then stimulated for 6 days with SARS-CoV-2 peptide pools or with IAV, and T cell proliferation was assessed based on CFSE dilution, using the gating strategy shown in **Fig. S4A**. 85% of donors showed CD4^+^ proliferative responses to S1 at their 1^st^ visit, 65% in response to N and 60% to IAV. These responses persisted for up to 35 weeks with a minimal decay of 0.3-0.5%/week, or a t1/2 of ∼100 weeks (**Fig. 4A**). CD8^+^ T cell proliferative responses were detected in 80% of donors at the first timepoint in response to S1, 75% to N and 100% to IAV. These responses decayed more rapidly than CD4^+^ T cell proliferative responses, decaying at a rate of 1-2% per week (**Fig. 4B**). T cell proliferative responses were also detected in response to S2, M and E in both CD4^+^ T cells and CD8^+^ T cells and decayed at similar rates (**Figs. 4C, 4D, S4B, S4C**). The data further show that within each donor, proliferative responses to S are most often dominated by CD4^+^ responses, whereas responses to N peptides are approximately equally divided between CD4^+^ and CD8^+^ T cells. In contrast, proliferative responses to IAV were dominated by CD8^+^ T cells (**Fig. 4E**). Altogether, the data show that while there is expansion of memory CD4^+^ and CD8^+^ T cells upon re-exposure to SARS-CoV-2 peptides, CD8^+^ proliferative responses are less durable than CD4^+^ proliferative responses, and responses to S are dominated by CD4^+^ T cells.

**Fig. 4.**
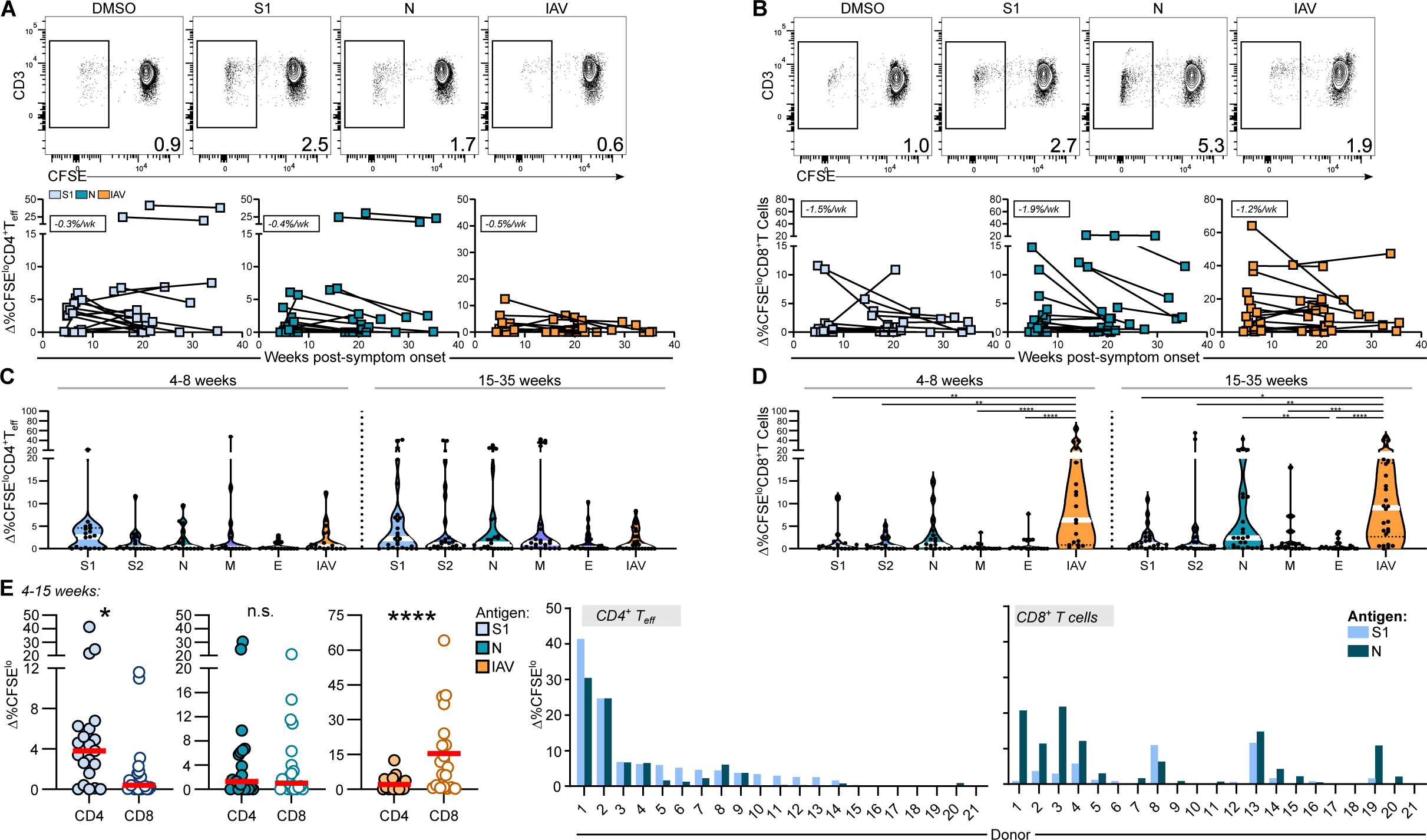
Robust CD4^+^ and CD8^+^ T cell proliferative responses to SARS-CoV-2 up to 35 weeks PSO. **(A)** Representative flow cytometry plots and longitudinal analyses of background subtracted values (Δ) show the frequency of CFSE^lo^CD4^+^ T cells after stimulation with the indicated antigen. Boxed numbers in each panel show the average rate of decay between time points for each response. **(B)** Representative flow plots and longitudinal analyses of background subtracted values (Δ) showing the frequency of CFSE^lo^CD8^+^ T cells. **(C)** Violin plot showing the median (white line) %CFSE^lo^CD4^+^ T cells binned by time PSO. **(D)** Violin plot showing the median (white line) %CFSE^lo^CD8^+^ T cells binned by time PSO. **(A)**-**(D)** N=24. **(C)** and **(D)** Comparisons were made by Dunn’s multiple comparisons test. **(E)** Comparison of the %CFSE^lo^ CD4^+^ or CD8^+^ T cells in response to each antigen using pairwise two-tailed Wilcoxon test. Right 2 graphs show the %CFSE^lo^ CD4^+^ or CD8^+^ T cells stimulated with S1 or N peptide pools, by individual donors. Wk = week. *P≤0.05, **P≤0.01, ***P≤0.001, ****P≤0.0001.

### Persistence of anti-SARS-CoV-2 antibodies in the plasma up to 35 weeks PSO

Antibodies are often an important correlate of protective immunity, and evidence suggests that this is the case in protection from COVID-19 (*27, 28, 41-44*). Therefore, we assayed plasma levels of S, RBD and N-specific IgG, IgA and IgM, for 19 study subjects for whom these samples were available. We found that most COVID-19 convalescent subjects (95%) mounted an IgG response against Spike trimer at the first visit, 79% against RBD and 53% against N, whereas IgA responses were lower, and few subjects demonstrated an IgM response to SARS-COV-2 S, RBD or N, which is not unexpected given that our first time point is several weeks past initial infection. IgG responses against Spike trimer were stable and persisted for up to 35 weeks, decaying at ∼ 0.50%/week (t1/2=100 weeks) respectively, whereas IgG responses against RBD and N were estimated to decay at rates of 1.8%/week (t1/2=28 weeks) and 1.1%/week (t1/2=45 weeks) respectively (**Fig. 5A**).

**Fig. 5.**
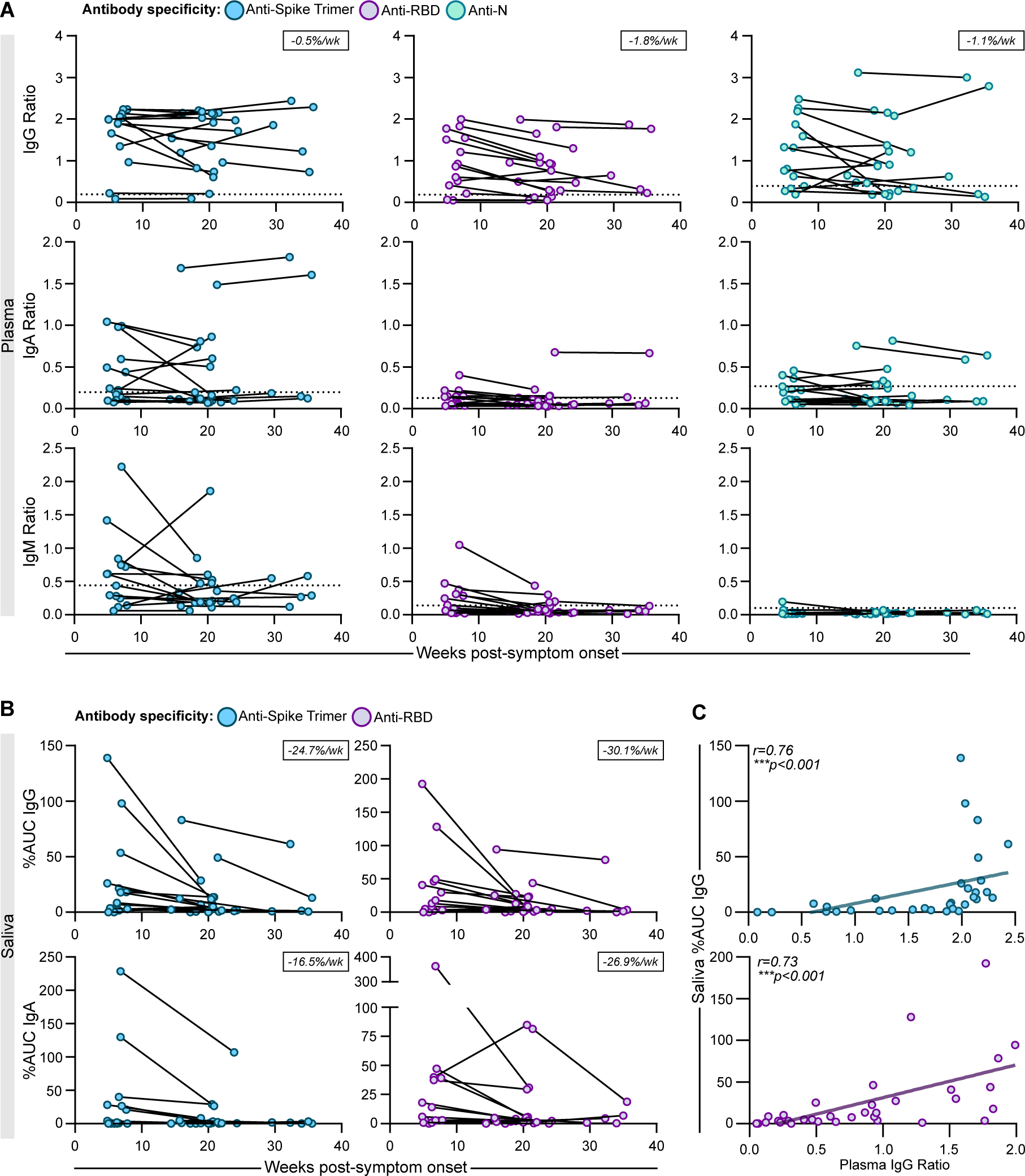
Persistence of anti-SARS-CoV-2 antibodies in the plasma up to 35 weeks post-symptom onset. Longitudinal analysis of the relative ratio (to a synthetic standard, see Methods) of IgG (top row), IgA (middle row) and IgM (bottom row) to Spike trimer, RBD or N in the plasma of COVID-19 convalescent individuals over time (N=19). Boxed numbers in the IgG panels show the average rate of decay between time points for each response. Dotted line indicates 3 standard deviations over the mean of the log negative control distribution (see Methods). **(B)** Longitudinal analysis of the %AUC IgG (top row) and IgA (bottom row) to Spike trimer or RBD in the saliva of COVID-19 convalescent individuals over time (N=19). **(C)** Correlation between plasma and salivary IgG to Spike trimer (top) or RBD (bottom). %AUC refers to area under the curve for experimental sample over AUC of positive control. Both timepoints for each donor are shown (N=17 per timepoint). Analysis was performed by Spearman correlation. Wk = week. *P≤0.05, **P≤0.01, ***P≤0.001, ***P≤0.0001.

We also measured antibodies in saliva and found that IgG responses in saliva decayed more rapidly than in the plasma, decreasing at 24-30% per week overall (**Fig 5B**). IgA responses also decayed rapidly in saliva, decaying at a rate of 17%/week against Spike trimer (t1/2=3.0 weeks) and 27 %/week against RBD (t1/2=1.9 weeks) (**Fig. 5B**). Plasma IgG strongly correlated with salivary IgG levels against both Spike trimer (r=0.76) and RBD (r=0.73) (**Fig. 5C**). 1/19 donors did not mount any antibody response to any of the Ags assayed, albeit T cell responses were detected in this individual. Taken together, our data show that the vast majority of COVID-19 convalescent subjects mounted an antibody response in the plasma and saliva to at least one SARS-CoV-2 protein, with little decay of anti-S plasma IgG over the course of the study, and a strong correlation between salivary and plasma IgG responses, as previously reported (*12*).

### Circulating Tfh and CD4^+^ Teffector responses correlate with plasma antibody levels

Tfh cells are important in the formation of memory B cells and long-lived antibody-producing plasma cells (*45*). While these cells typically reside in the germinal centers (GC) of lymphoid organs, cTfh cells are the peripheral counterparts of GC Tfh; cTfh are able to efficiently induce antibody secretion by B cells, contribute to antibody generation following vaccination and correlate with broadly neutralizing antibody responses against some viruses (*46–48*). Here, we identified SARS-CoV-2-specific cTfh by ICC staining as IL-2-producing CD45RA^-^ CXCR5^+^CCR7^+^ CD4^+^ T cells with gating shown in **Fig. S1A**. At first visit, 63% of subjects had a cTfh response to S1, 71% to N and 92% to IAV. While cTfh responses to S1 were detected out to 35 weeks PSO, decaying at a rate of 2%/week (t1/2=22 weeks), there were large variations and fluctuations in N- and IAV-specific cTfh responses over time (**Fig. 6A**), albeit this may reflect the low frequency of responses. cTfh responses, again at low frequencies, could also be detected in 71% of subjects against S2, 57% to M, and 48% to E. As expected, based on whole IAV having the entire complement of Ags, the frequency of IL-2^+^ cTfh was greatest after IAV stimulation (**Figs. 6B, S5**).

**Fig. 6.**
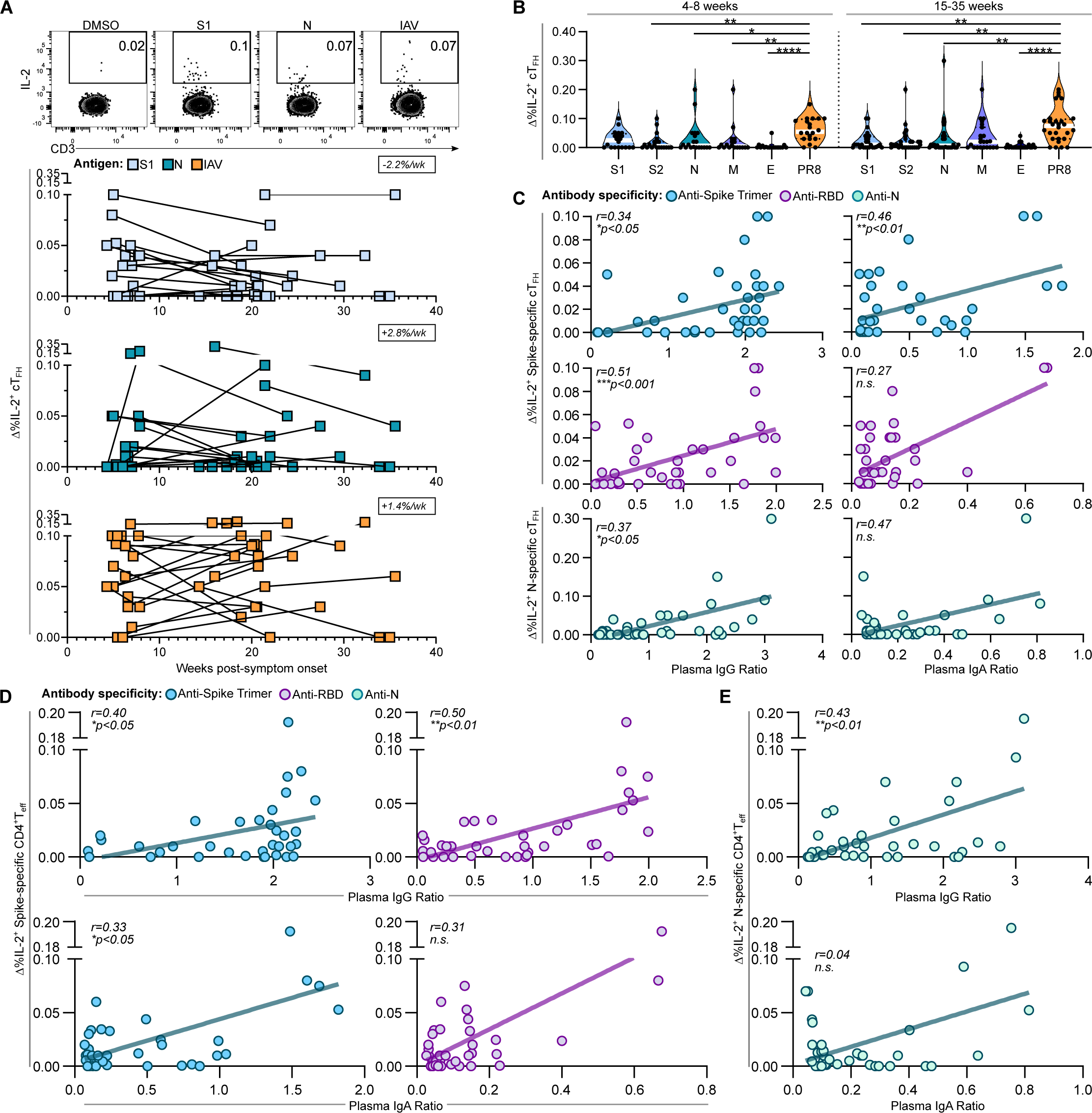
Circulating Tfh and CD4^+^ Teff responses correlate with plasma antibody levels. **(A)** Representative flow plots and longitudinal analyses of background subtracted values (Δ) show the frequency of cTfh expressing IL-2 to the indicated antigen (N=24). Boxed numbers in each panel show the average rate of decay between time points for each response **(B)** Violin plots show the median frequency (white line) of cTfh expressing IL-2 to each antigen binned by time PSO (4-8 weeks or 15-35 weeks). Comparisons were made by Dunn’s multiplex comparisons test (N=24). **(C)** Correlation between S-specific IL-2^+^cTfh and anti-Spike trimer or anti-RBD IgG in the plasma (top and middle row), and correlation between N-specific IL-2^+^cTfh and anti-N IgG in the plasma (bottom row). **(D)** Correlation between S-specific IL-2^+^CD4^+^ Teff and Spike-/RBD-specific IgG (top) or IgA (bottom) in the plasma. Boxed numbers in each panel show the average rate of decay between time points for each response **(E)** Correlation between N-specific IL-2^+^CD4^+^ Teff and N-specific IgG (top) or IgA (bottom). **(C)**-**(D)** Correlation analysis in was performed by Spearman correlation, with each plot depicting both timepoints for each donor (N=19 per timepoint). Wk = week. **P≤0.05, **P≤0.01, ***P≤0.001, ***P≤0.0001.

There was a positive correlation between the frequency of S-specific IL-2^+^ cTfh and plasma IgG responses mounted against Spike trimer and Spike RBD, as well as plasma anti-Spike trimer IgA (**Fig. 6C**). However, there was no correlation between Tfh and anti-RBD IgA in the plasma. Likewise, the frequency of N-specific IL-2^+^cTfh significantly correlated with anti-N IgG in the plasma, but not with anti-N IgA (r=0.47), although there was a positive trend (**Fig. 6D**). Similarly, IL-2^+^ S-specific CD4^+^ T effector cell (Teff) responses (defined as the non-Tfh IL-2-producing cells), significantly correlated with both plasma anti-Spike trimer IgG, IgA, and anti-RBD IgG. Again, there was no correlation with plasma anti-RBD IgA (**Fig. 6D**). Analysis of IL-2^+^ N-specific CD4^+^ Teff responses also showed significant correlation with anti-N IgG in the plasma (r=0.43, p<0.01), and likewise did not correlate with levels of anti-N IgA (**Fig. 6E**). In sum, cTfh responses can be detected in response to SARS-CoV-2 S, N and M peptide pools for most of the subjects out to almost 9 months PSO, and although these responses are of low frequency, both the SARS-CoV-2-specific cTfh and CD4^+^ Teff responses correlated with plasma antibody titers.

### Analysis of cytokine secretion by SARS-CoV-2 convalescent PBMCs compared to IAV

We next used a quantitative multiplex bead array to determine the levels of 13 cytokines/cytolytic molecules released into the supernatant after 48h stimulation of PBMC with SARS-CoV-2 peptide pools or with IAV. Cytokines produced in response to S and N included IFN-γ, IL-2 and TNF and were detectable up to 35 weeks PSO. IL-2, IL-10 and TNF induction by IAV were stable up to 35 weeks, as was IL-10 induction by N peptides. The TNF recall response diminished most rapidly, with a reduction of 4.5%/week in response to S, and 6%/week in response to N (**Fig. 7A**). IL-4 and IL-17a were detected at low levels in some donors with no consistent trend observed over time (**Fig. S6**). Additionally, IL-6, a pro-inflammatory cytokine, was substantially upregulated at the first visit in 96% of subjects when stimulated with S peptide pools, and in 75% of subjects with N, compared to 62.5% of subjects when stimulated with IAV and was maintained up to 35 weeks PSO (**Fig. 7A**). In sum, while both SARS-CoV-2 and IAV recall responses are characterized by a Th1 profile, the majority of cytokines measured in response to SARS-CoV-2 Ags declined over time, with the exception of IL-6 and IL-10 responses, whereas the responses to IAV were more stable.

**Fig. 7.**
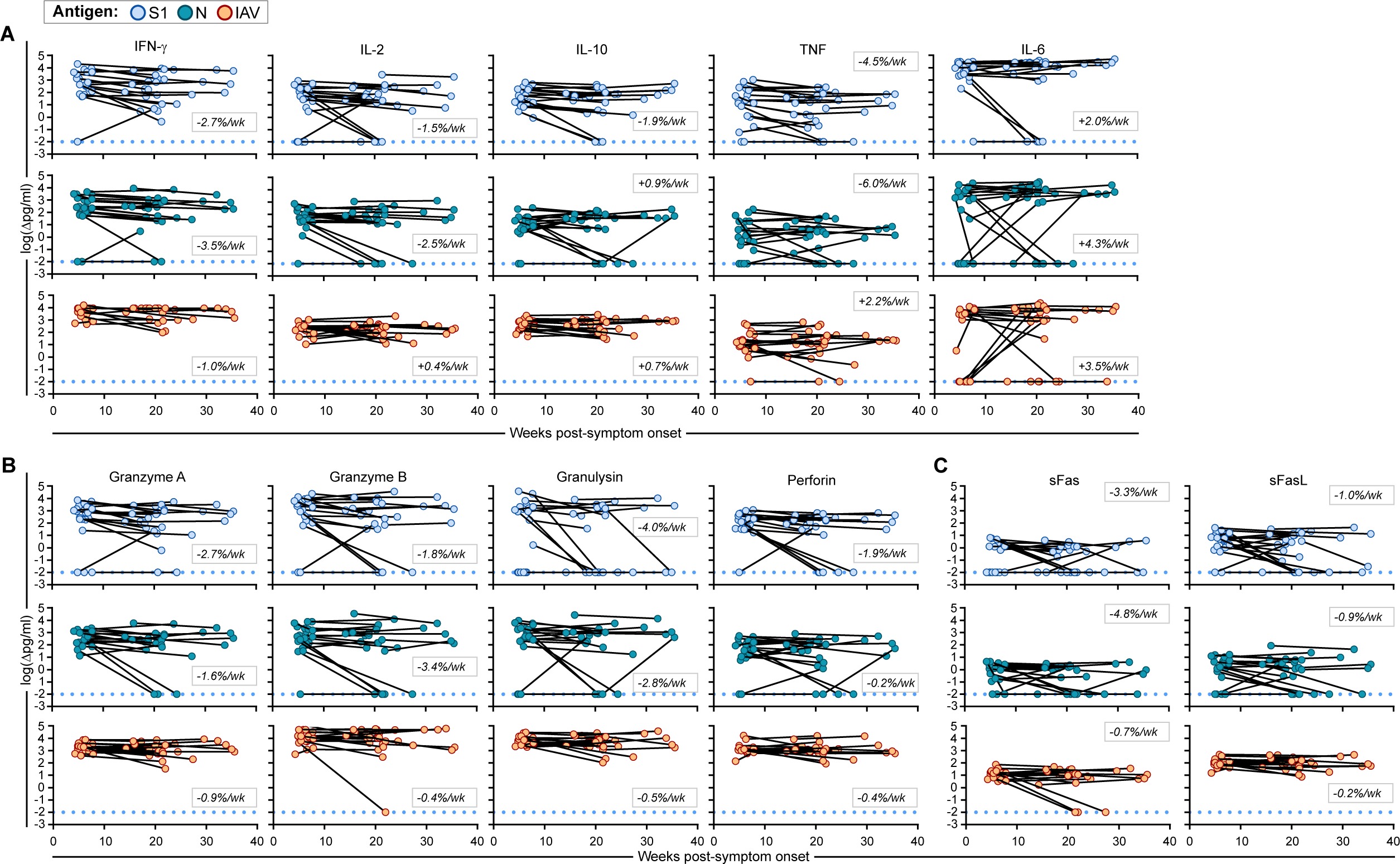
Analysis of cytokine secretion by SARS-CoV-2 convalescent PBMCs compared to IAV. Cytokines in the supernatant of COVID-19 convalescent PBMC cultures were measured using a multiplex bead array after 48h stimulation with S1 or N peptide pools, or IAV (N=24). Graphs show longitudinal analysis of background subtracted values (Δ) of secreted **(A)** IFN-γ, IL-2, IL-10, TNF, IL-6, **(B)** granzyme A, granzyme B, granulysin, perforin and **(C)** sFas, sFasL. Dotted blue line indicates the limit of detection. Wk = week. Boxed numbers in each panel show the average rate of decay between time points for each response

Molecules associated with cytotoxicity, including granzyme A, granzyme B, granulysin and perforin, were detected in response to both S and N peptide pools. However, the response to S and N showed a wider range of responses and more rapid decay in general compared to IAV (**Fig. 7B**). sFas and sFasL were both upregulated in 100% of subjects in response to IAV with minimal decay over time. In contrast, sFas was upregulated in only 58% of subjects in response to S and in 75% of subjects to N, both decaying more rapidly than IAV. sFasL was upregulated in 88% of subjects to S1 and 83% of subjects to N, decaying at a rate of 0.9%/week (**Fig. 7C**).

### SARS-CoV-2 convalescent PBMCs exhibit an altered secreted cytokine profile compared to IAV

As we observed differences in cytokine upregulation and durability in response to SARS-CoV-2 Ag compared to IAV, we next sought to characterize differences in the cytokine profile induced by each viral Ag. To this end, we calculated fold-change in response to S peptide or IAV and performed hierarchical agglomerative clustering on log-transformed data. Values across donors were normalized per cytokine as described in the Methods. At 4-8 weeks PSO, we found that S1 and IAV stimulated samples did not completely cluster based on Ag stimulation, while at 15-35 weeks PSO, samples clustered almost completely by type of Ag (**Fig. 8A, B**). Analysis of the clusters revealed that while most donors upregulated IFN-γ, Granzyme A, Granzyme B, and IL-10 (associated with typical antiviral responses), there was a cluster of study subjects who had noticeably diminished expression of Granzyme A, Granzyme B and IL-10 (“Module 1”). S1-specific responses also exhibited lower expression of sFas, sFasL, Granulysin and perforin (“Module 2”) compared to IAV within the same set of donors. TNF and IL-2 were similarly upregulated between S1- and IAV-stimulated samples, while IL-17A and IL-4 were not broadly upregulated in response to either Ag. (**Fig. 8A**). We observed that these two cytokine modules were conserved, and more apparent, at 15-35 weeks PSO (**Fig. 8B**).

**Fig. 8.**
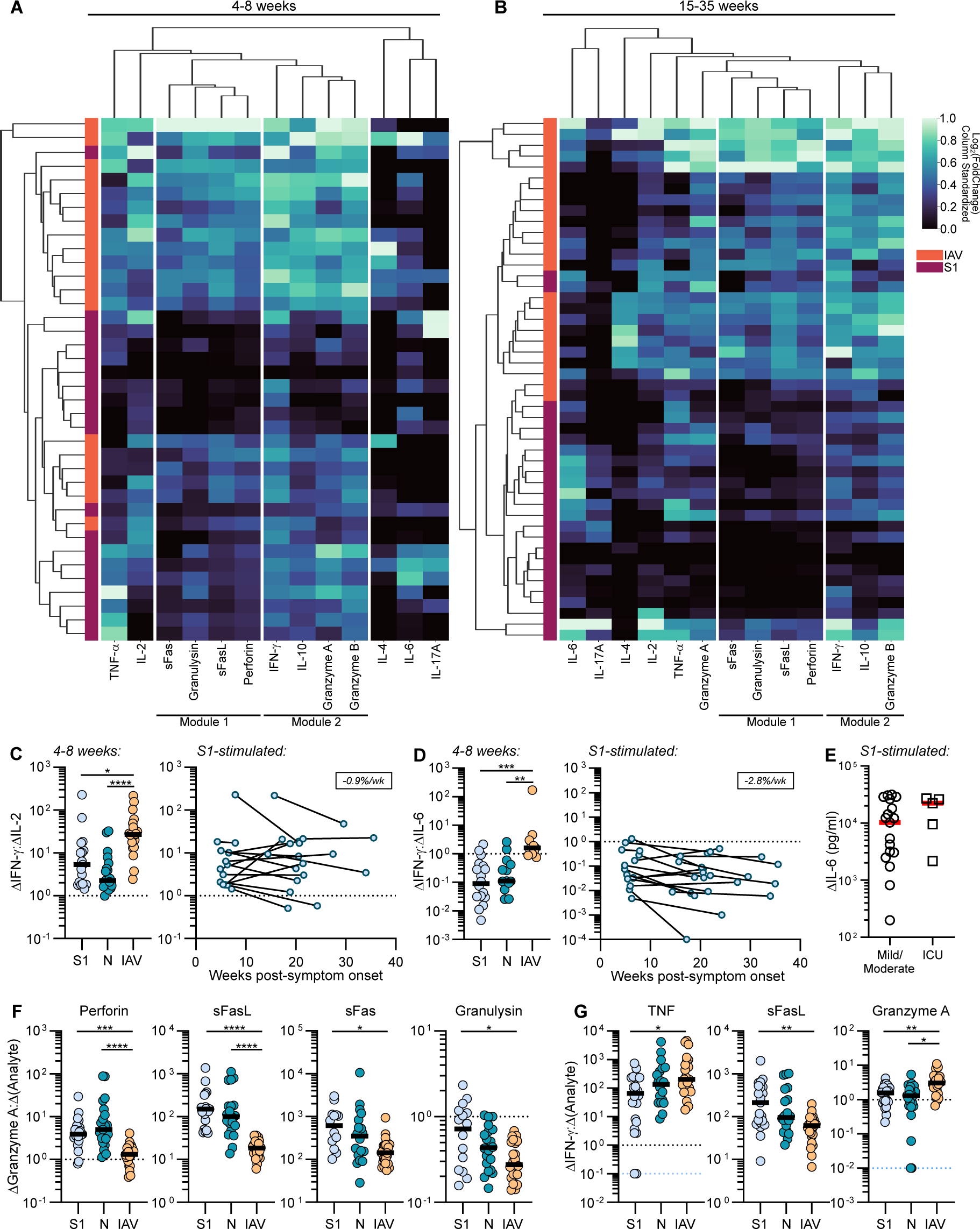
SARS-CoV-2 convalescent PBMCs exhibit an altered secreted cytokine profile compared to IAV. Agglomerative hierarchical clustering analysis of the fold change in secreted cytokines was performed on log-transformed data, comparing responses to S1 peptide pool or IAV in each donor at **(A)** 4-8 weeks and **(B)** 15-35 weeks. Each row represents 1 donor, and each column represents 1 cytokine. Values across donors for each cytokine were standardized by min/max normalization. **(C)** Ratio of IFN-γ:IL-2 at the early convalescent phase 4-8 weeks PSO (right), and over time to S1 peptide pools (left), N=20. **(D)** Ratio of IFN-γ:IL-6 at the early convalescent phase 4-8 weeks PSO (right), and over time to S1 peptide pools (left), N=19. Boxed numbers in the panels in **(C)** and **(D)** show the average rate of decay between time points for each response. **(E)** Comparison of IL-6 secretion between mild/moderate and ICU subjects at the first visit in response to S1 peptide pools (N=24). **(F)** Ratio of Granzyme A to perforin, sFasL, sFas and granulysin at the first visit (N=24). **(G)** Ratio of IFN-γ to TNF, sFasL and Granzyme A at the first visit (N=24). **(C)**-**(G)** Comparisons were made by Dunn’s multiple comparisons test. Wk = week. * P≤0.05, **P≤0.01, ***P≤0.001, ****P≤0.0001. Dotted black line indicates 1:1 ratio. Dotted blue line indicates values at the limit of detection. “Δ” indicates values with background subtracted.

As the overall response to whole IAV is higher than the response to each of the SARS-CoV-2 peptide pools analyzed, to determine if there was indeed a different profile of cytokine response to SARS-CoV-2 Ags compared to IAV, we determined the ratio IFN-γ to IL-2 or IL-6 within each donor for IAV or SARS-COV-2-specific responses. Accordingly, we observed a reduced IFN-γ: IL-2 ratio in response to S and N peptide pools, compared to IAV stimulation. Furthermore, this reduced ratio of IFN-γ: IL-2 was retained over time, recapitulating our observations based on ICC analysis (**Figs. 2B, 8C, S7A**). The data also revealed a greatly reduced IFN-γ:IL-6 ratio in response to SARS-CoV-2 Ags compared to IAV. Similarly, this altered IFN-γ:IL-6 ratio persisted up to at least 35 weeks PSO (**Fig. 8D, S7B**).

To investigate whether the profile of cytotoxic molecules was also distinct between SARS-CoV-2-specific responses and IAV-specific responses independently of the overall magnitude of the response, we analyzed the proportion of cytotoxic molecules secreted in response to S, N or IAV, relative to levels of Granzyme A and IFN-γ within each donor for each Ag. S-specific responses resulted in a lower level of Perforin, sFas, sFasL and granulysin, relative to Granzyme A, compared to IAV whereas N-specific responses differed from IAV-specific responses in perforin and sFasL only (**Fig. 8F**). Relative to the level of IFN-γ in the cultures, IAV specific responses showed differences in TNF, sFasL and Granzyme A relative to SARS-CoV-2 specific responses (**Fig. 8G**). Therefore, after normalizing to account for different overall level of response, data from the cytokine/cytotoxic molecule secretion assay demonstrate an altered antiviral profile in response to SARS-CoV-2 compared to IAV, which is less IFN-γ dominated, more pro-inflammatory based on IL-6, with an altered cytotoxic profile that might indicate different anti-viral mechanisms.

## Discussion

Here we investigated the longevity of antibody and T cell responses to SARS-CoV-2 by comparing two memory time points in a cohort of 24 SARS-CoV-2 convalescent subjects, the majority of whom were non-hospitalized. Based on ICC staining, we detected CD4^+^ Teff responses to S and N in 80-95% of subjects, cTfh responses in 60-70% of subjects and CD8^+^ T cell responses in only 30-50% of study participants. Proliferation assays similarly showed a higher proportion of CD4^+^ over CD8^+^ T cell responses in SARS-CoV-2 recovered subjects. Although there was a wide range in the magnitude of responses, all subjects showed a response to at least one SARS-CoV-2 Ag in at least one T cell assay. However, the phenotype of the SARS-CoV-2 specific T cells differed from that of memory responses to IAV within the same donor, with a lower CD8^+^: CD4^+^ T cell ratio and a lower ratio of IFN-γ producing to IL-2 producing CD4^+^ T cells based on proliferation and ICC data. Analysis of secreted molecules confirmed the finding of a lower ratio of IFN-γ to IL-2 in SARS-CoV-2-specific responses, as well as demonstrating a decreased IFN-γ: IL-6 ratio and an altered profile of cytotoxic molecules compared to IAV-specific responses. Antibody responses to SARS-CoV-2 were detected in 95% of subjects and there was a correlation between saliva and plasma IgG. Our findings demonstrate that most individuals in a cohort of recovered, mainly mild cases have detectable T and antibody responses to SARS-CoV-2 out to almost 9 months post-infection. These findings are consistent with recent studies showing 80-85% protection from re-infection with SARS-CoV-2 in the under 65 age group with up to 7 months of follow-up (*49–52*).

Plasma IgG responses to S, although variable between individuals, were remarkably stable over the course of the study decaying at 0.5% per week (t1/2=23 months), whereas anti-RBD IgG response had a t1/2 of around 6 months. S and RBD-specific antibody responses in saliva fell off more rapidly. Nonetheless, there was a correlation between the plasma and saliva anti-S and anti-RBD IgG levels, as previously noted (*12*). Given that anti-SARS-CoV-2 antibody titers in the saliva are lower than in the plasma at all time points, it is possible that the more rapid decline in saliva may reflect levels of antibody in the saliva that drop below the limit of detection of the assay. There was also a correlation between levels of plasma IgG and IgA and the cTfh and the Teff response. The longevity of IgG responses to S and the slightly more rapid decay of anti-RBD responses, as well as the more transient IgA and IgM responses, are similar to that reported in earlier studies (*12, 22, 34*).

On average we found that CD4^+^ T cells specific for S or N decayed at a rate of about 2% per week (t1/2 5-6 months) between the two time points based on IL-2 secretion, or at a rate of 2-3%/week (t1/2 4-5 months) based on activation markers (4-1BB, OX40). Tfh responses (IL-2^+^CCR7^+^CXCR5^+^) also decayed at 2-3% per week. CD8^+^ T cell responses detected based on IFN-γ production were considerably weaker than CD4^+^ T cell responses and declined at 1-2% per week (t1/2 ∼7 months). A caveat to these estimates is that the T cell frequencies based on ICC were low, potentially rendering them less quantitative. Nonetheless, our results are quite similar to those of Dan et al. (*34*), who reported a half-life of 6 months for CD8^+^ T cell responses and 3 months for CD4^+^ T cell responses, by analyzing paired samples based on activation markers. SARS-CoV-2 specific proliferative responses of CD4^+^ T cells were more stable between time point one and two than the proliferative responses of CD8^+^ T cells, perhaps reflecting the higher proportion of Tcm in the CD4^+^ population and a higher proportion of Temra in the CD8^+^ T cell populations. Bilich, et al. reported stable CD8^+^ T cell responses and increasing CD4^+^ T cell responses over 2 time points (median 40 and 159 days) based on ICC or ELIspot assays, in a cohort of SARS-CoV-2 convalescent subjects, most with mild disease (*36*). However, the study of Bilich et al. (*36*) was focused on a series of defined epitopes, whereas the study of Dan et al. (*34*) and the present study used overlapping peptide pools covering the full Ag sequences. Thus, although the overall T cell response to SARS-CoV-2 declines over time, it is possible that subsets of epitope-specific T cells form longer lived subsets, which displace other T cells over time.

The responses to IAV in our study were about 2-3 times more stable than the response to SARS-CoV-2 within the same subjects. As boosting is known to increase the duration of T cell immunity (*53*), this finding likely reflects that the IAV-specific memory has been boosted over a lifetime of exposure or vaccination, whereas the response to SARS-CoV-2 represents the memory response to a single infection. On the other hand, the primary response to the Yellow fever vaccine strain YF17D in humans gives rise to long-lived HLA-A2/NS4b-specific CD8^+^ T cells with a half-life of 493 days (*54*). Thus, the longevity of T cell responses likely reflects the stimulation history as well as the particular virus and epitope studied.

Both IAV-specific and SARS-CoV-2-specific T cells showed a mixture of memory phenotypes, with SARS-CoV-2 S-specific CD4^+^ T cell responses showing a higher proportion of Tcm (CD45RA^-^ CD27^+^) than IAV-specific responses, whereas IAV-specific responses showed a higher proportion of effector memory T cells (CD45RA^-^CD27^-^) at both time points. Curiously both IAV and SARS-CoV-2 -specific T cells included a proportion of CD45RA^+^ CD27^+^ T cells, a phenotype normally associated with naïve T cells. As these cells were detected based on Ag-specific cytokine production they seem unlikely to be naïve but might represent incompletely differentiated memory T cells. We also included overnight restimulation with P/I in our assays, which is thought to preferentially induce cytokine production by memory T cells in short term cultures (*55*). The rate of decay of these responses was similar to the rate of decay of IAV-specific responses, and likely represents the overall stability of T cell memory. This decline in response to P/I is unlikely due to assay effects, as for each study subject samples from the 2 time points were thawed and assayed at the same time and the number of HLA-DR^+^ APC was similar between time points overall.

Based on intracellular cytokine staining, S1, S2, N and M specific CD4^+^ T cells were detected in most donors, whereas responses to E were quite rare. More than 80% of these Ag-specific CD4^+^ T cells produced only 1 cytokine, with IL-2 producing T cells most frequent, followed by IFN-γ and then TNF. Within the same samples, IFN-γ producing cells predominated over IL-2 producing CD4^+^ T cells following stimulation of the PBMC with IAV. Moreover, within each donor there was a higher frequency of CD8^+^ over CD4^+^ IAV-specific T cells, whereas SARS-CoV-2 S-specific responses showed a predominance of CD4^+^ over CD8^+^ responses, as previously noted (*8, 11, 16, 19*). IAV stimulation involves whole virus, and therefore includes multiple viral proteins as compared to our analysis of SARS-CoV-2 responses based on separate peptide pools for the Ag studied. Consistently, the overall level of activation of IAV-specific T cells at time point 1 and 2 was higher than T cell responses to S1 peptide pools based on activation markers, including OX40, 4-1BB and PD-1. Nonetheless, the different proportion of cytokines within each response suggests a different phenotype in the SARS-CoV-2 S- or N-specific versus IAV-specific CD4^+^ response that is not explained by the different magnitude of the response. IAV-specific responses were also slightly more multifunctional than SARS-CoV-2-specific responses. This might be due to IAV-specific memory representing a lifetime of potential exposures, whereas the response to SARS-CoV-2 represents a primary infection; whether this will change upon boosting remains to be determined.

In a previous study, in which about half the study subjects were hospitalized or ICU subjects, we also observed that the most frequent SARS-CoV-2-specific T cells produced IL-2 (*19*). However, in that study, TNF-producing CD4^+^ T cells were also more frequent than IFN-γ producing cells (*19*), whereas in the present study, TNF-producing CD4^+^ T cells were less frequent. It is possible that the higher proportion of hospitalized cases in the previous study impacted the levels of TNF-producing cells observed (*19*). As the median time of sampling in the previous study 37 days (range 27-90) was not that different from time point one of the current study, median 45 days (range 30-154 days), we think it is unlikely that time since infection impacts the proportion of TNF-producing cells, albeit this can’t be ruled out.

Based on the multiplex bead assay, the most predominant cytokines secreted in response to S and N were IFN-γ and IL-6. Unsupervised clustering of the 13 secreted molecules largely segregated SARS-CoV-2-specific from IAV-specific responses, particularly at the later time point. IAV specific responses were higher overall, and showed a higher cytotoxic profile, likely due to the stronger stimulation from whole virus compared to individual peptide pools. However, by looking at the ratio of cytokines produced within each donor, we observed an altered cytokine profile for SARS-CoV-2-specific as compared to IAV-specific responses, findings that recapitulated the results from ICC analysis. Notably, within each donor, the ratio of IFN-γ: IL-2 was lower for S- and N-specific T cell responses compared to IAV-specific responses. We also noted a lower level of IFN-γ: IL-6 in S- and N-stimulated cultures compared to IAV-stimulated cultures. IL-6 can be secreted by a variety of cells including monocytes and macrophages as well as T cells (*56*). Whether IL-6 was produced directly by activated CD4^+^ or CD8^+^ T cells, or by other cells responding to activated T cells or their cytokines cannot be deduced from these data. However, as the same PBMC preparation was used to investigate IAV- and SARS-CoV-2-specific recall responses for each subject, it appears that differences in IL-6 production are Ag-specific and therefore attributed to differences in the T cells rather than APC. IL-6 can limit IFN-γ production by T cells (*56*) and has been shown to limit cytotoxicity in severe COVID (*57*). Thus, the higher Ag-specific SARS-CoV-2-dependent production of IL-6 at the memory phase may represent a fundamental difference between SARS-CoV-2 and IAV-specific T cell responses, contributing to the overall phenotype.

Here we have used 3 different assays to characterize T cell responses to SARS-CoV-2. The flow cytometry-based ICC assay, which detects Ag-specific responses based on 18h of restimulation, has the advantage of providing single cell data on cytokine production and other markers, at a time point before cells start to divide, thus providing an estimate of Ag-specific T cells in the blood immediately ex vivo. However, the ICC assay has the disadvantage that the frequency of responses we measured were quite low in many of the subjects, particularly for the CD8^+^ T cell responses, potentially rendering the analysis less quantitative. To compensate for this, we also used a proliferation assay, which confirmed the lower CD8^+^: CD4^+^ ratio for SARS-CoV-2 S-specific responses compared to IAV-specific responses,observed by ICC. A caveat to the proliferation assay, however, is that the long period of T cell expansion could amplify weak cross reactive T cell responses to seasonal CoV, as pointed out in a recent study (*58*). We also measured cytokines produced in the culture supernatants after 48hr of Ag stimulation, using a quantitative commercial bead-based assay. A limitation of this assay is that it does not indicate which cell produced the cytokine in question, albeit the cytokines were specifically detected in response to Ag stimulation. Despite these limitations, the altered cytokine profile detected in SARS-CoV-2 specific as compared to IAV-specific T cells by ICC was recapitulated by the multiplex cytokine assays and the latter may be useful in future for larger studies, due to its ease of high throughput analysis.

In sum, we report that SARS-CoV-2 -specific T cell responses are largely dominated by IL-2 producing CD4^+^ T cells, with a less frequent CD8^+^ T cell response and a potentially altered cytotoxic mechanism. The higher production of IL-6 in SARS-CoV-2-specific compared to IAV-specific recall responses may contribute to this altered phenotype. Although a limitation of our study is that the IAV-specific responses measured here represent a lifetime of exposure to IAV and stimulation with whole virus, whereas SARS-CoV-2-specific responses represent the response to a new infection and peptide pools representing a subset of Ags, the data suggest that SARS-CoV-2 specific T cell responses are distinct from the typical response to the respiratory pathogen IAV.

## Materials and Methods

### Study design

This study was designed to investigate the persistence and immunophenotype of T cell and antibody responses to SARS-CoV-2 in subjects who had recovered from PCR-confirmed COVID-19 and to compare this phenotype to IAV-specific memory in the same subjects. Each subject donated blood and saliva at two time points, the first between 30-154 days post-symptom onset (PSO), median 45 days and the second between 55-249 days PSO, median 145 days. Saliva and plasma were used for analysis of antibody responses, whereas PBMC were stimulated with SARS-CoV-2 peptide pools or whole IAV, and assayed using three complementary assays: a 16-parameter flow cytometry panel, in which cytokine production was used to identify Ag-specific T cells after 18h of stimulation; a proliferation assay in which dilution of CFSE by CD4^+^CD3^+^ or CD8^+^ CD3^+^ was measured after 6 days of stimulation, and a commercial multiplex bead array to analyze 13 secreted cytokines and cytotoxic molecules in the supernatant after 48h of stimulation.

### Human subjects and sample preparation

Individuals who had recovered from COVID-19 as confirmed by positive nasopharyngeal COVID-19 PCR upon presentation, were recruited to RISC-CoV through participating hospitals in the Toronto Invasive Bacterial Diseases Network, with informed consent, to donate blood and saliva, as approved by the University of Toronto Research Ethics Board (REB protocol number 00027673 to THW). All human subjects research was done in compliance with the Declaration of Helsinki. Whole blood was collected from healthy human donors by venipuncture and plasma separated. Saliva was collected in salivette tubes, as described (*12*). Saliva and plasma aliquots were stored frozen at -80°C until use. Peripheral blood mononuclear cells (PBMC) were isolated by density gradient centrifugation using Ficoll-Paque PLUS (GE Healthcare). PBMC were stored at -150°C in AIM-V (Gibco) 10% DMSO until use.

### T cell stimulation assay for intracellular cytokine detection

Cryopreserved PBMCs were thawed at 37°C, washed twice with PBS and cultured in complete media (RPMI 1640 supplemented with 10% FBS, 2-mercaptoethanol, sodium pyruvate, penicillin, streptomycin and non-essential amino acids (Gibco) at 37°C with 5% CO2. 2x10^6^ PBMCs were plated per well in 96-well round bottom plates for 18h with 1 μg/ml of SARS-CoV-2 S1, S2, N, M, or E peptide pools (JPT PepMix, S1/S2: 90% purity, N/M/E:70% Purity) or 100 hemagglutination units (HAU)/ml of live IAV PR8/34. PBMCs were cultured with equimolar DMSO as a negative control. GolgiStop (BD Biosciences, San Jose, CA) containing monensin was added in the last 6h of the culture. As a positive control, 50 ng/ml PMA (Sigma-Aldrich), 1 μg/ml ionomycin (Sigma-Aldrich), GolgiStop was added to PBMCs cultured with complete media in the last 6h of culture.

### Intracellular cytokine staining

After culture, PBMCs were washed with PBS containing 2% FBS (FACS buffer). Cells were first stained with anti-human CCR7 [clone G043H7 (BioLegend, San Diego, CA)] at 37°C for 10 min, followed by staining with Fixable Viability Dye eFluor™ 506 (eBiosciences, Thermofisher, Mississauga, Ontario) to discern viable cells, anti-human CD3 [clone UCHT1 (BioLegend)], CXCR5 [clone J252D4 (BioLegend)], 4-1BB [clone 4B4-1 (BioLegend)], HLA-DR [clone L243 (BioLegend)], CD4 [clone SK3 (BD)], CD27 [clone L128 (BD)], CD8 [clone Sk1 (eBiosciences)], PD-1 [clone EH12.2H7 (BioLegend)], OX40 [clone Ber-ACT35 (BioLegend)], CD69 [clone FN50 (eBiosciences)] and CD45RA [clone HI100 (BD)] for 20 min at 4°C. Cells were washed twice with FACS buffer, then fixed with BD Cytofix/Cytoperm buffer (BD Biosciences) for 20 min at 4°C. Following fixation and permeabilization, cells were washed twice with 1X BD Perm/Wash buffer (BD Biosciences) and stained with anti-human IFN-γ [clone 4S.B3 (Biolegend)], TNF [clone Mab11 (BioLegend)] and IL-2 [clone MQ1-17H12 (eBiosciences)] for 15 min at 4°C. Samples were washed twice, then resuspended in FACS buffer and acquired on the BD LSRFortessa X-20 flow cytometer using FACSDiva software.

### Multiplex cytokine bead assay

1x10^6^ PBMCs were seeded per well in 96-well round bottom plates with 1 μg/ml each of SARS-CoV-2 S1, S2, N, M, or E peptide pools (JPT PepMix), or 100 HAU/ml live IAV. PBMCs were cultured with equimolar DMSO as a negative control. Cell culture supernatants were collected after 48h of incubation at 37°C and stored at -80°C. Cytokines in the supernatants were measured using the Human CD8/NK Cytokine Panel (13-plex) LEGENDplex kit (Biolegend) with capture reagents specific for IL-2, IL-4, IL-10, IL-6, IL-17A, TNF, sFas, sFasL, IFN-γ, granzyme A, granzyme B, perforin and granulysin. The assay was performed as per the manufacturer’s instructions using a V-bottom plate. Samples were acquired on the BD LSRFortessa X-20 flow cytometer.

### CFSE proliferation assay

PBMC were labeled with 10 μM of CFSE (Thermo Fisher Scientific) in PBS for 3 min at room temperature. Excess CFSE dye was quenched by adding 10x volume of cold complete media and incubated on ice for 5 min. Cells were then washed twice and resuspended in complete media and plated at 4x10^5^ cells/well in 96-well round bottom plates. CFSE-labelled PBMCs were incubated with 1 μg/ml of SARS-CoV-2 S1, S2, N, M, or E peptide pools (JPT PepMix), 100 HAU/ml live PR8, or equimolar DMSO (negative control) for 6 days at 37°C. On day 6, cells were washed with FACS buffer, and stained with Fixable Viability Dye eFluor™ 506 (eBiosciences) to discern viable cells, anti-human CD3 [clone UCHT1 (BioLegend)], CD4 [clone RPA-T4 (BD)], CD8 [clone SK1 (eBiosciences)], CD45RA [clone HI100 (BD)] and CD27 [clone L128 (BD)] for 20 min at 4°C. Cells were washed twice with FACS buffer, then fixed with BD Cytofix for 20 min at 4°C. Samples were washed twice, then resuspended in FACS buffer and acquired on the BD LSRFortessa X-20 flow cytometer using FACSDiva software.

### ELISA assays for detecting antibodies in plasma

An automated chemiluminescent ELISA assay was used to analyze the levels of IgG, IgA and IgM antibodies to the spike trimer, its RBD, and the nucleocapsid, essentially as in (*12*)with the following modifications. The nucleocapsid antigen was produced in CHO (Chinese Hamster Ovary) cells and was a kind gift from Dr. Yves Durocher, National Research Council of Canada (NRC). For IgG, the secondary antibody was an IgG-HRP fusion, provided by the NRC. A standard curve of the VHH72 monobody (*59*) fused to human IgG1 Fc domain (also from the NRC) was generated for calibrating the anti-spike and anti-RBD IgG response. All other antigens, detection reagents and calibration reagents were as previously described (*12*). The analysis also proceeded largely as in (*12*), with the following exceptions. Blanks were not subtracted from the chemiluminescence raw values of the samples, and the raw values were normalized to a blank-subtracted point in the linear range of the calibration standard curve (for Spike and RBD, the reference point was 0.0156 ug/ml and for N, 0.0625 ug/ml). The final results are represented as a “relative ratio” to this calibration standard point. To define the cutoff for positive antibody calls for each antigen for IgG, 3 standard deviations from the mean of the log negative control distribution from 20 different runs collected over 4 months was used as a cut-off to define positivity in each individual assay. For IgA, negatives from 2 different runs over one month and for IgM negatives from 3 runs over 2 months were used for this. In all cases, this corresponds to <2% False Positive Rate (FPR) assessment, based on Receiver Operating Characteristic Curves. The selected cutoff for positivity was drawn as a dashed line on the figure.

### ELISA assays for detecting antibodies in saliva

The expression, purification and biotinylation of the SARS-CoV-2 RBD and spike ectodomain were performed as previously described (*12, 60*) Enzyme-linked immunosorbent assays to detect Spike and RBD-specific IgG and IgA antibodies in saliva were completed as described in reference (*12*). To analyze the data, raw OD450 measurements obtained from PBS-coated wells corresponding to each sample diluted at 1/5 (“background signal”) was subtracted from readings obtained from Ag-coated wells at each of the three dilutions described (1:5, 1:10, 1:20). For each plate, a sample of pooled saliva from COVID-19 acute and convalescent subjects was plated at 1/5 with no Ag (PBS control), as well as with Ags at 1:5, 1:10 and 1:20. The area under the curve was calculated based on the background subtracted values from all three dilutions for each sample and for the pooled sample of positive control saliva run on each plate. Each sample within a given plate was then normalized to the pooled positive control saliva for that particular plate and expressed as a percentage.

### Data and Statistical analysis

Flow cytometry data were analyzed using FlowJo v10.7.1 (BD Biosciences). Multiplex cytokine bead data were analyzed using the LEGENDplex™ Data Analysis Software Suite (BioLegend). Statistical analyses were performed using GraphPad Prism v9.1.1. “Δ” in figure labels represents values for which background signal has been subtracted. “Background signal” is defined as the frequency of cells expressing the parameter being analyzed, or concentration of an analyte, in negative control wells. In ICC and proliferation data, the response was considered positive if the response to an Ag was 10% higher than the background signal. In multiplex data, the response was considered positive if the value was above the limit of detection, which is established using a standard curve for each analyte. Pairwise comparisons were made by a two-tailed Wilcoxon test, or non-parametric Dunn’s multiple comparisons test. Non-pairwise comparisons were made by Mann-Whitney test. Correlation analyses were performed by computing the Spearman correlation coefficient. For longitudinal data, the percent change over time and t1/2 were calculated after linear regression was performed for each donor. We report the average percent change per week and average t1/2. Cluster analysis for multiplex data was performed by first calculating fold-change in response to an Ag, and then performing hierarchical agglomerative clustering on log-transformed data. Values across donors were standardized by min/max normalization such that for each analyte, the minimum is subtracted, then divided by its maximum. These data were visualized using the Seaborn data visualization library for Python (https://joss.theoj.org/papers/10.21105/joss.03021).

Supplementary Materials

Fig. S1. Gating strategy and cytokine production by CD4+ T cells to SARS-CoV-2 S2, M and E, and P/I

Fig. S2. Distribution of memory subsets in SARS-CoV-2-specific and IAV-specific CD4+ T cells

Fig. S3. Distribution of memory subsets in SARS-CoV-2-specific and IAV-specific CD8+ T cells

Fig. S4. Gating strategy and T cell proliferation to S2, M and E peptide pools

Fig. S5. Circulating Tfh responses to SARS-CoV-2 S2, M and E peptide pools

Fig. S6. IL-4 and IL-17A secretion by SARS-CoV-2 convalescent PBMCs

Fig. S7. Altered cytokine profile in SARS-CoV-2 convalescent PBMC persists over time

Table S1. Participant characteristics

## Data Availability

Data and materials availability: All data files, including Flow cytometry FCS plots, are available from the authors upon request.

## Acknowledgments

We thank Birinder Ghumman for technical assistance, Andrew Law for assistance with data analysis and Seaborn implementation, Nathalie Simard and Janine Charron for assistance with flow cytometry, Yves Durocher at the NRC for antigens, calibration standards and secondary antibodies, members of the Gingras laboratory, Adrian Pasculescu and the Network Biology Collaborative Centre for assistance with sample intake and antibody data generation and analysis, Thierry Mallevaey for critical reading of the manuscript, and all the study subjects for participating and donating samples to make this study possible.

## Author contributions

J.C.L and T.H.W. designed experiments and conceptualized the study; M.A.O, S. M. and A.J.M. recruited study subjects. G.C. and M.G. conducted blood draws and saliva acquisition and assisted with patient scheduling. M.G., Y.Y.F. and J.C.L processed PBMC. M.A.O. and A.J.M. recruited study subjects. J.C.L. conducted all T cell experiments and analyzed data; B.I., L.W. and J.G. analyzed antibody responses in saliva; B.R., K.C. and A.C.G. analyzed antibody responses in plasma. JR and ZL prepared biotinylated antigens for saliva ELISA assays. JCL and THW wrote the manuscript which was reviewed and edited by all contributing authors.

## Data and materials availability

All data files, including Flow cytometry FCS plots, are available from the authors upon request.

## Supplementary Figure Legends

**Fig. S1.**
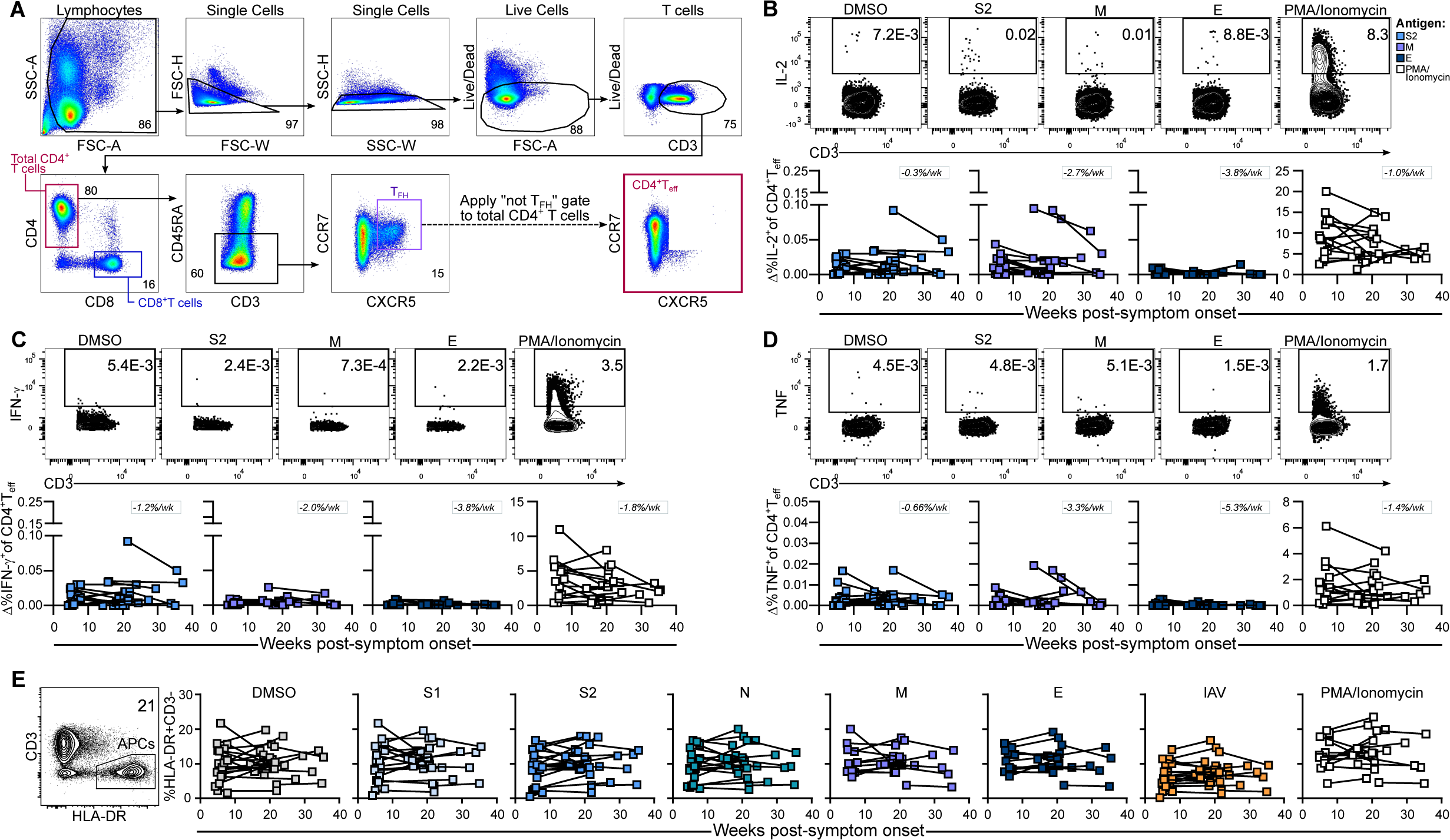
Gating strategy and cytokine production by CD4^+^ T cells to SARS-CoV-2 S2, M and E, and P/I. **(A)** Representative gating strategy for cTfh, non-cTfh CD4^+^ Teff and CD8^+^ T cells. Representative flow plots and longitudinal analyses of background subtracted values (Δ) show the frequency of CD4^+^ T cells expressing **(B)** IL-2, **(C)** IFN-γ, and **(D)** TNF in response to S2, M and E peptide pools, or PMA/Ionomycin positive control. (E) Representative gating strategy for antigen-presenting cells (APCs). Graphs depict the %HLA-DR^+^CD3^-^ cells over time in response to the indicated antigen, or to PMA/Ionomycin control. **(B)**-**(E)** N=24. Wk = week.

**Fig. S2.**
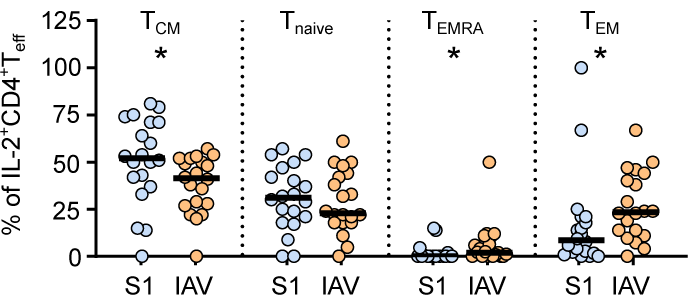
Distribution of memory subsets in SARS-CoV-2-specific and IAV-specific CD4^+^ T cells. Graph shows each memory subset as a proportion of IL-2^+^CD4^+^ T cells at the first visit. Pairwise comparisons of the proportion of each memory subset between S1 and IAV-specific CD4^+^ T cells were performed by two-tailed Wilcoxon test. N=20. *P≤0.05.

**Fig. S3.**
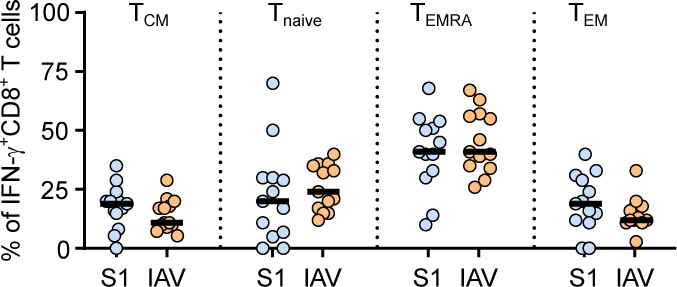
Distribution of memory subsets in SARS-CoV-2-specific and IAV-specific CD8^+^ T cells. Graph shows each memory subset as a proportion of IFN-γ^+^CD8^+^ T cells at the first visit. Pairwise comparisons of the proportion of each memory subset between S1 and IAV-specific CD8^+^ T cells were performed by two-tailed Wilcoxon test. N=13.

**Fig. S4.**
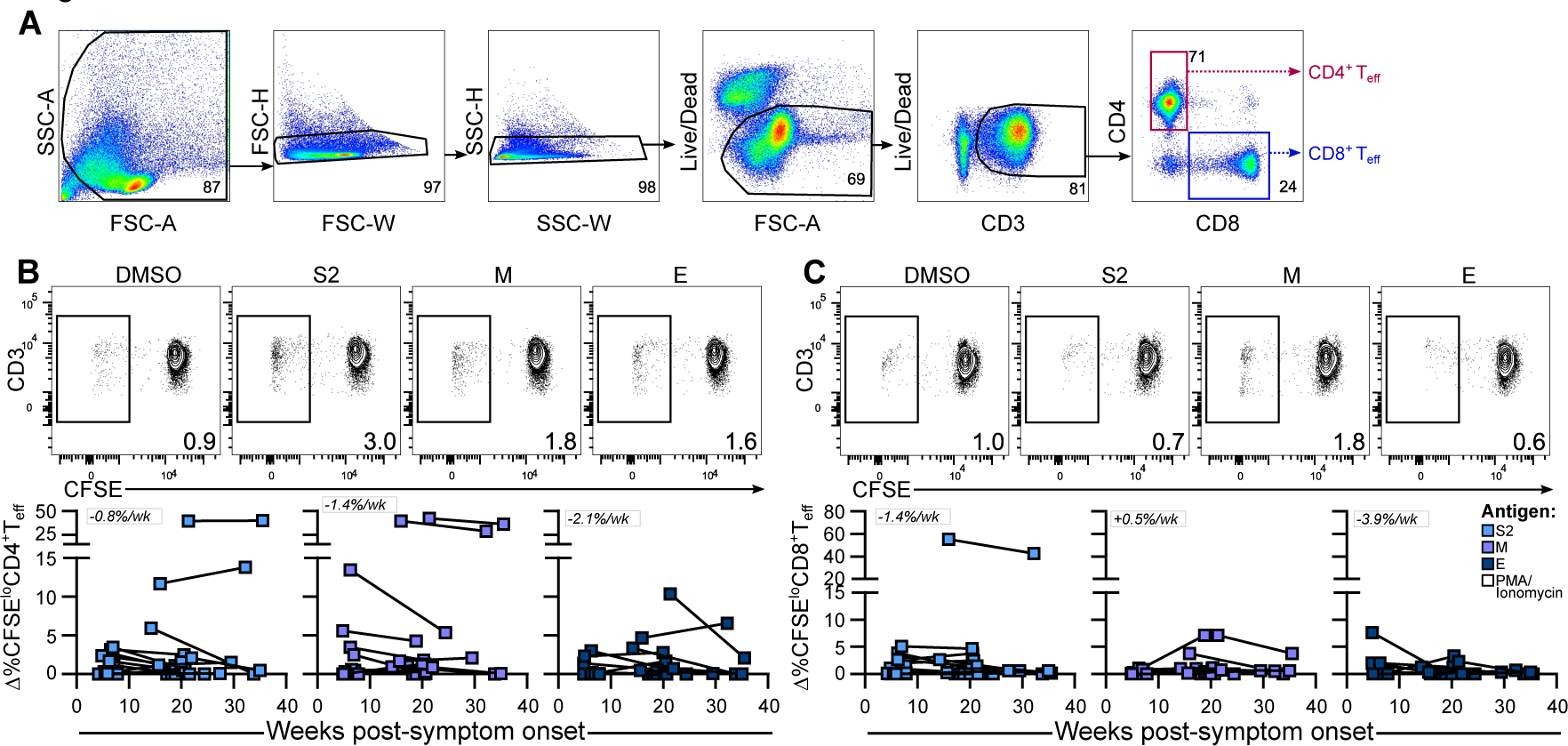
Gating strategy and T cell proliferation to S2, M and E peptide pools. **(A)** Representative gating strategy for CD4^+^ and CD8^+^ T cells. Representative flow plots and longitudinal analyses of background subtracted values (Δ) show the frequency of **(A)** CFSE^lo^CD4^+^ T cells and **(B)** CFSE^lo^CD8^+^ T cells after stimulation with S2, M or E peptide pools. N= 20. Wk = week.

**Fig. S5.**
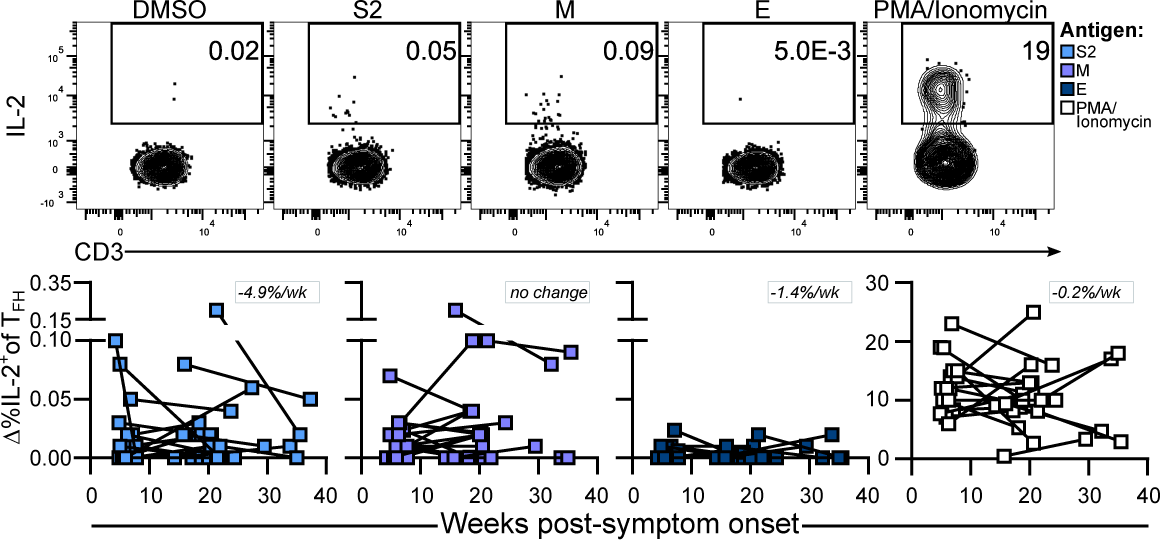
Circulating Tfh responses to SARS-CoV-2 S2, M and E peptide pools. Representative flow plots and longitudinal analyses of background subtracted values (Δ) show the frequency of cTfh cells expressing IL-2 in response to S2, M and E peptide pools, or PMA/Ionomycin positive control. N=24. Wk = week.

**Fig. S6.**
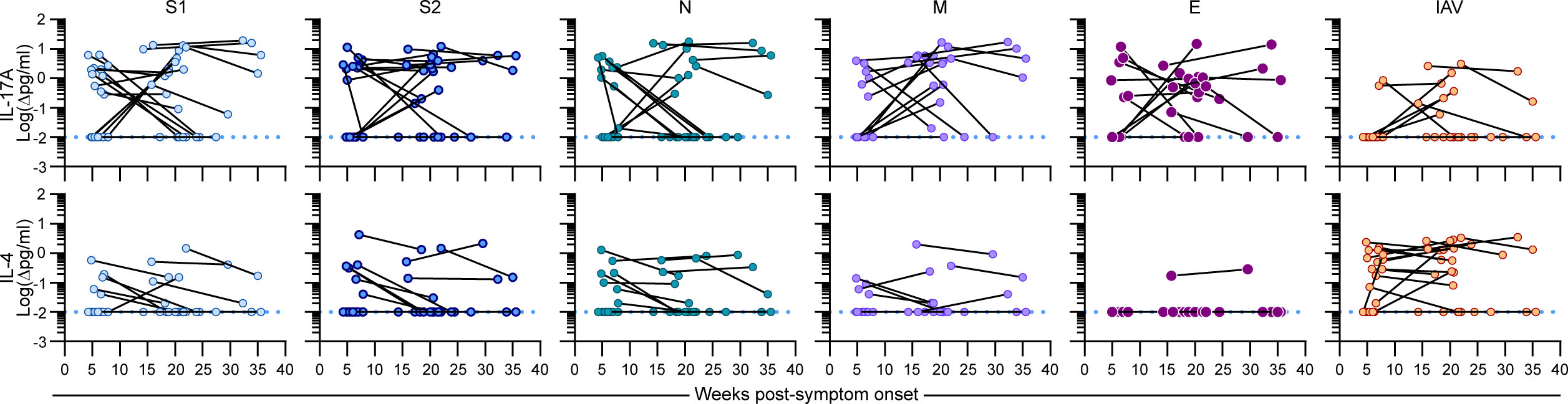
IL-4 and IL-17A secretion by SARS-CoV-2 convalescent PBMCs. Cytokines in the supernatant of COVID-19 convalescent PBMC cultures were measured using a multiplex bead array after 48h stimulation with S1, S2, N, M or E peptide pools, or whole IAV. Graphs show longitudinal analysis of background subtracted values (Δ) of secreted IL-17A (top row) and IL-4 (bottom row). N=24. Dotted blue line indicates the limit of detection.

**Fig. S7.**
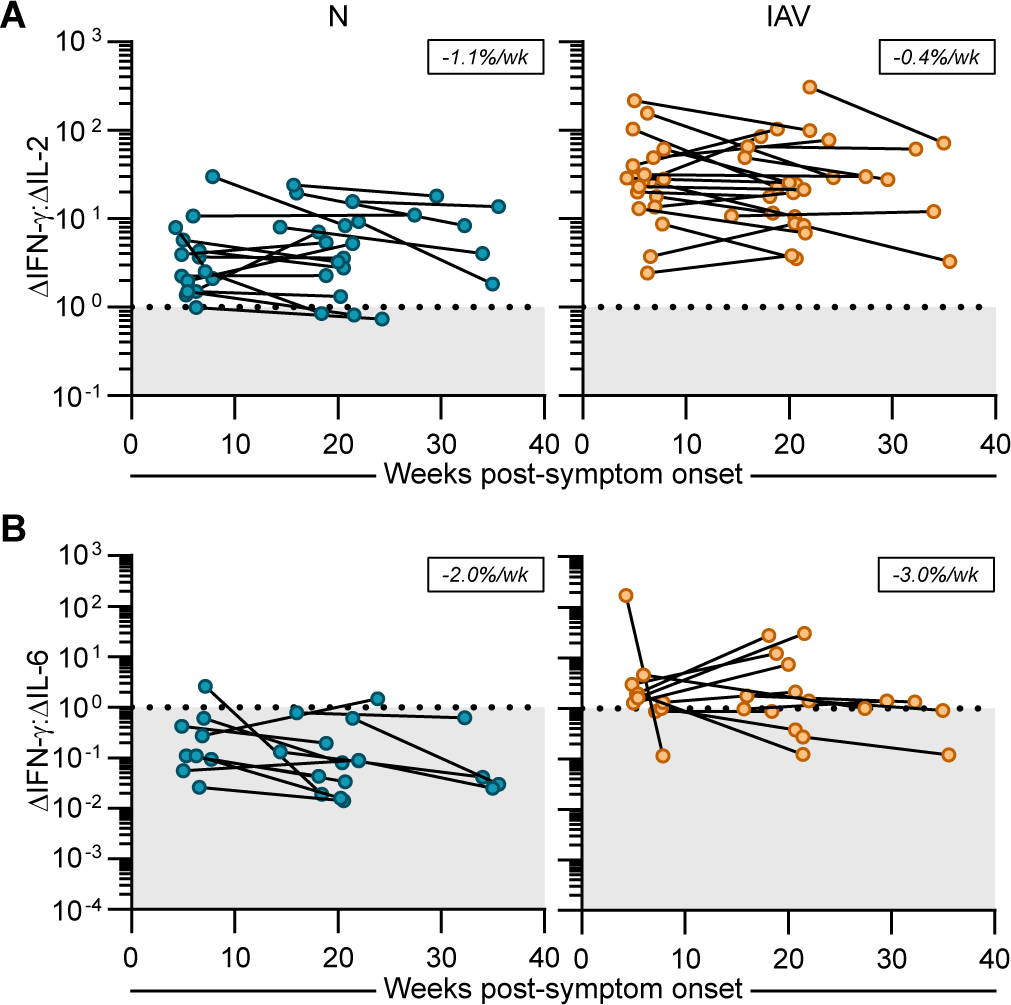
Altered cytokine profile in SARS-CoV-2 convalescent PBMC persists over time. **(A)** Ratio of secreted IFN-γ:IL-2 over time in response to N peptide pools (left, N=20) and IAV (right, N=24). **(B)** Ratio of secreted IFN-γ:IL-6 over time in response to N peptide pools (left, N=17) and IAV (right, N=19). “Δ” indicates values with background subtracted. Wk = week. Dotted black line indicates 1:1 ratio.

**Table S1.**
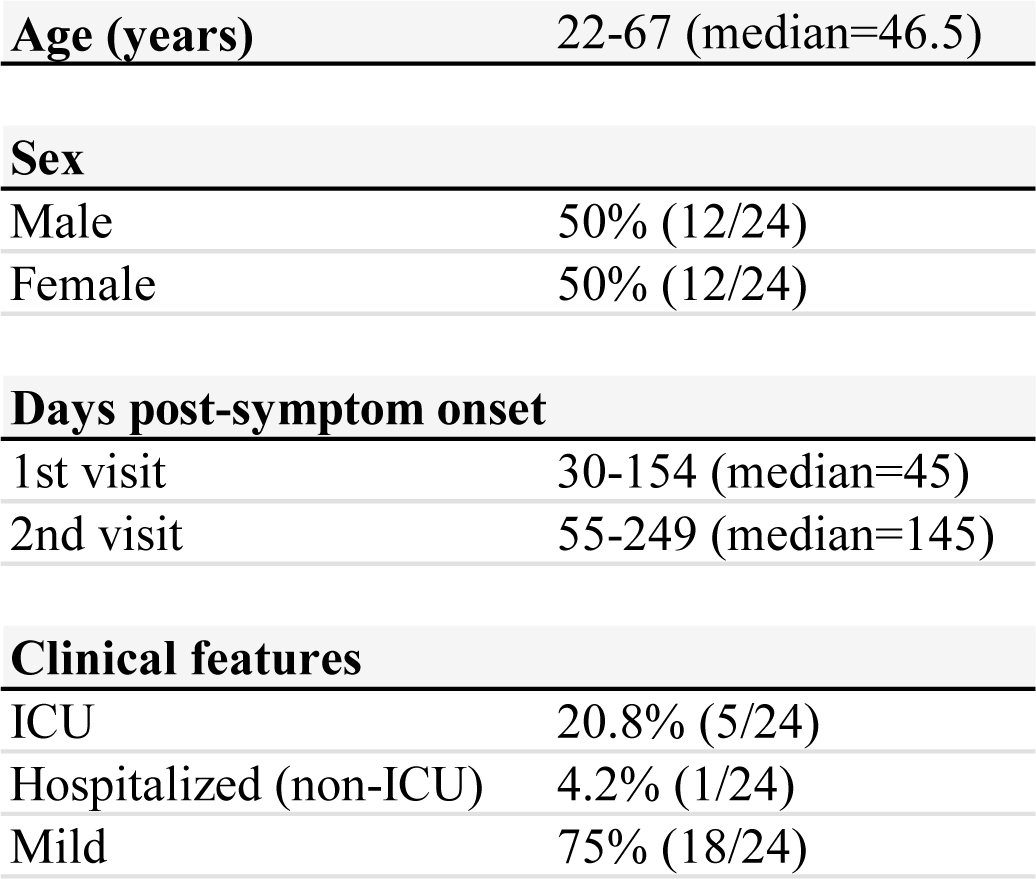
Participant characteristics.

|  |  |
| --- | --- |
| **Age (years)** | 22-67 (median=46.5) |
| **Sex** |  |
| Male | 50% (12/24) |
| Female | 50% (12/24) |
| **Days post-symptom onset** |  |
| 1st visit | 30-154 (median=45) |
| 2nd visit | 55-249 (median=145) |
| **Clinical features** |  |
| ICU | 20.8% (5/24) |
| Hospitalized (non-ICU) | 4.2% (1/24) |
| Mild | 75% (18/24) |

